# Identification and Validation of New Molecular Subtypes within the Early and Late Mild Cognitive Impairment Stages of Alzheimer’s Disease

**DOI:** 10.1101/2023.04.06.23288268

**Authors:** Bowei Li, Shashank Yadav, Shu Zhou, Yuheng Du, Leyuan Qian, Zachary Karas, Yueyang Zhang, Li Shen, Jenny J. Lee, Badri N. Vardarajan, Annie J. Lee, Lana X Garmire, Alzheimer’s Disease Neuroimaging Initiative (ADNI)

**Affiliations:** Department of Computational Medicine and Bioinformatics, the University of Michigan. Ann Arbor, MI, USA. 48105; Department of Biostatistics, the University of Michigan. Ann Arbor, MI, USA. 48109; Department of Statistics, the University of Michigan. Ann Arbor, MI, USA. 48105; Department of Biostatistics, Epidemiology and Informatics, University of Pennsylvania, Philadelphia, PA, USA 19104; Department of Data Science, Ewha Womans University, Seoul 03760, Republic of Korea; The Department of Neurology, Vagelos College of Physicians and Surgeons, Columbia University and the New York Presbyterian Hospital, New York, NY, USA. 10032; The Gertrude H. Sergievsky Center, Columbia University, New York, NY, USA. 10032; Taub Institute for Research on Alzheimer’s Disease and the Aging Brain, Columbia University, New York, NY, USA. 10032

**Keywords:** Alzheimer’s disease, Mild cognitive impairment, Multi-omics integration, metabolomics, transcriptomics, Similarity Network Fusion

## Abstract

Alzheimer’s disease (AD) is a heterogeneous neurodegenerative condition. This study identifies clinically relevant new molecular subtypes of the early and late mild cognitive impairment (EMCI and LMCI) stages of AD from 401 patients in the ADNI consortium. Metabolomics and peripheral blood mononuclear cell (PBMC) transcriptomics data are integrated using Similarity Network Fusion (SNF), resulting in two molecular subtypes within EMCI (EMCI-1 and EMCI-2) and LMCI (LMCI-1 and LMCI-2), respectively. Metabolomics, rather than gene expression profiling, is the major contributor to subtyping. Subtype-specific differences in metabolite levels and gene expression correlate with AD pathophysiology, supported by cognitive scores, pseudo-trajectory analysis, and longitudinal dementia diagnoses. The new molecular subtypes are further validated through the EFIGA dataset, and present a clear trend of increase in protein biomarkers including p-Tau181, p-Tau217, A*β40*, and A*β42*. These refined subtypes represent a step toward more personalized therapeutic strategies targeting early disease stages before AD diagnosis.

## 1. INTRODUCTION

Alzheimer’s Disease (AD) is a terminal neurodegenerative disease with irreversible cognitive impairment ^1^. Patients with AD experience memory loss, lose motor function, have trouble eating and swallowing, and struggle to communicate with their family members ^2,3^. In 2010, roughly 35.6 million people worldwide lived with AD, which is projected to increase to 115.4 million by the year 2050 due to the increasing average lifespan ^4^. Effective therapeutics would improve the lives of millions of patients and their families and lighten the economic burden on societies ^5^. However, there is still no cure for AD ^6^ despite the significant effort to develop treatments ^7^.

There are three general stages along the AD continuum: first preclinical, then mild cognitive impairment (MCI), and finally dementia. Since identifying the MCI stage of AD in the late 1990s ^8^, researchers have recognized this intermediate stage as having more potential for therapeutic interventions than the dementia stage ^9^. Many lifestyle and pharmaceutical interventions have been tested at the MCI stage, with some degrees of success ^10–12^. However, further improvement in the MCI population by therapeutic interventions is still elusive, largely because of the characteristic heterogeneity of AD ^13^. Thus, developing a more granular understanding of the MCI stage of AD could provide new directions for further research and elucidate better targets for clinical applications ^14^.

Metabolomics is an emerging omics field and provides a temporally specific and global view of an individual’s ongoing biological processes associated with disease phenotypes ^15,16^. Metabolomics is sensitive to transient biological processes, and transcriptomics data have uncovered more targeted biomarkers in cancer studies as compared to other omics platforms ^17^. While there is a growing body of metabolomics research for AD ^18^, the multi-omics AD study to integrate metabolomics to other omics data types is currently lacking ^19^. The other omics approach, such as Extracellular RNAs, have emerged to be a promising biomarker for AD presymptomatic identification as well ^20^. Therefore, in this study, we aim to use the multi-omics approach to better understand the heterogeneity of AD and to identify coherent subtypes ^19,21^.

In particular, we hypothesize that integrating transcriptomics and metabolomics data will yield more subtypes in MCI, an important pre-clinical stage of the AD continuum. Using data from MCI patients in the Alzheimer’s Disease Neuroimaging Initiative (ADNI) cohort, we applied an unsupervised multi-omics integration method called Similarity Network Fusion (SNF) ^22^ and identified two subtypes in EMCI (EMCI-1 and EMCI-2) and LMCI (LMCI-1 and LMCI-2) groups, respectively. We confirmed these subtypes by various approaches: (1) we compared them with cognitively normal (CN) and AD patients to illuminate the characteristics of MCI in AD progression through different stages; (2) we performed pseudo-time trajectory analysis using the clinical information to validate the gradual change of subtypes’ severities; (3) we conducted the survival analysis to demonstrate the trends of onset time of AD among these groups; (4) we built label transferring models based on the EMCI vs LMCI group and subgroup labels in ADNI cohort and validated that them in the Estudio Familiar de Influencia Genetica en Alzheimer (EFIGA) dataset.

## 2. METHODS

### 2.1 ADNI Datasets

The ADNI consortium was launched in 2004^23^. This consortium began as a public-private collaborative effort across many countries. There have been multiple phases of the ADNI project, starting with ADNI-1, followed by ADNI-GO, then ADNI-2, and ADNI-3. The ADNI database includes clinical data, cognitive assessments, genomics and metabolomics data, MRI images, PET images, and biospecimens ^24^. In our analyses, we used transcriptomics and metabolomics data. There are 681 patients with both metabolomics and transcriptomics data from blood samples. Among them, 401 patients are classified as either EMCI (201) or LMCI (200), as shown in **Table 1**.

**Table 1.** Descriptive statistics for the subsample of ADNI data used in the current study. This includes patients diagnosed as either EMCI or LMCI at baseline who have both transcriptomics data and CSF metabolomics data available (* - significant differences between the two groups (P value < .001)).

|  | No. of cases (n) | Age (mean) | Gender | Race (%) | ApoE-ε4 carrier |
| --- | --- | --- | --- | --- | --- |
| **Early Mild Cognitive Impairment** | 201 | 70.85 | 94 Females | White (92.0)<br>Black (3.5)<br>Asian (1.5)<br>Hawaiian (0.5)<br>More than one (2.5) | 64 carriers |
| **Late Mild Cognitive Impairment** | 200 | 73.54* | 76 Females | White (94.5)<br>Black (2.5)<br>Asian (2)<br>Hawaiian (0.5)<br>More than one (0.5) | 74 carriers |

### 2.2 Transcriptomics and metabolomics data preprocessing of ADNI dataset

The raw transcriptomics data from ADNI contain gene expression data from over 48,000 transcripts. We used the average expression values of multiple transcripts to represent each gene, resulting in 20,032 genes. We used median values among duplicate assays, followed by quantile normalization before downstream analysis.

For the metabolomics data from ADNI, we first combined P180 Kit data from ADNI-1 and ADNI-2/GO, obtained from previous reports ^25^. To correct the batch effects, the correction factor for each analyte was calculated by dividing its average on a given plate by its global average. Metabolites were then removed if their coefficient of variation across plates was greater than 20% or more than 40% of its measurements across all subjects were not available. We excluded 107 patients from our analyses who reportedly did not fast before the blood draw, to minimize the confounding effect due to the diet. We also consolidated data from the 36 blinded replicates by calculating the average of the blinded duplicates for each of these subjects. Lastly, we transformed the values into the log-2 scale, centered and scaled the data, and replaced any values that were >3.0 standard deviation away from the center in either direction with 3.0 and -3.0, following the original reports. Such procedures yield 172 metabolites in 1,572 patients across every stage of AD, among which 681 are classified as MCI.

### 2.3 Dimension reduction of transcriptomics and metabolomics data using autoencoders

Given the considerable differences in the total numbers of metabolomics and genes (172 vs. ∼20,000) in the ADNI dataset, we first used an autoencoder ^26^ to reduce the dimensionality of gene expression to 172, to match the metabolomics data. We set the bottleneck layer of the autoencoder for gene expression as 172. We then integrate the metabolomics data with a higher-order representation of the transcriptomics data from the autoencoder’s bottleneck layer. This integration of transcriptomics data and metabolomics data forms the basis for further analyses.

### 2.4 Subtype identification within EMCI and LMCI

This used SNF for integration between gene expression and metabolomics data ^22^. SNF first constructs sample similarity matrices for each omics data, and then aggregates these matrices in a non-linear fashion. We first calculated the Silhouette Coefficients (SC) and Calinski Harabasz (CH) Scores for cluster sizes ranging from 2 to 15 to find the optimal number of clusters to fit the data from EMCI and LMCI patients, separately. An SC ranges between -1 to 1 to determine how distinguishable the clusters are from one another, with values closer to 1 being more distinguishable.

### 2.5 Deconvolution of transcriptomics data by cell type

We performed deconvolution of the transcriptomics data from PBMC whole blood samples of the ADNI dataset to get the proportions of different cell types in leukocytes using the GEDIT package ^27^. Specifically, we deconvoluted 22 human hematopoietic cell phenotypes, including seven T-cell types, naive and memory B cells, plasma cells, two natural killer (NK) cell types and 10 myeloid subsets by using a reference dataset from one of GEDIT’s reference matrices, termed LM22 ^28^, with the abovementioned 22 cell types. After deconvolution, we preserve all the cell types with an average proportion greater than 3% for the later analysis (eg. differential expression).

### 2.6 Differential Gene Expression Analysis

With the newly defined subgroups and deconvoluted cell proportions in the ADNI dataset, we first performed differential expression (DE) analysis using the limma package in R among the subtypes of EMCI and LMCI, respectively. The subtypes in less severe disease status were set as the control, namely EMCI-1 and LMCI-1. We added the proportion levels of different blood cell types as the covariates of the DE analysis, in addition to the subtypes and other clinical covariates including age, race and gender. Genes with adjusted *P* value <.05 from the DE analysis were subject to additional thresholding of absolute fold change >1.3 were considered as the significant DE genes. We then performed gene set enrichment analysis (GSEA) on the DE genes to uncover the upregulated and downregulated pathways.

For metabolomics data, DE analysis was conducted using the limma package, with adjustment for clinical covariates age, race and gender, followed by pathway analysis using MetaboAnalyst 6.0 web tool ^29^. The thresholds for significant differential metabolites are adjusted *P* value <.05 and absolute fold change >1.2.

### 2.7 Contextualizing Patient Subtypes with Clinical Data

We clustered patients within the EMCI and LMCI groups using metabolomics and transcriptomics data and then checked whether these subsamples showed any significant differences in cognitive measures, which are relied upon to diagnose patients as healthy, MCI, or AD ^9^. The ADNI consortium includes data from a battery of cognitive tests, with key measures being the Mini-Mental State Exam (MMSE) and the Clinical Dementia Rating (CDR). We analyzed differences between the subtypes for these metrics and additionally investigated patients’ scores on the Functional Activities Questionnaire (FAQ), the Alzheimer’s Disease Assessment Scale (ADAS-11, ADAS-13, and ADAS-Q4), the Trail Making Test (TRABSCOR), the Delayed Recall Total score (LDELTOTAL), and subtests from the Rey Auditory Verbal Learning Test (RAVLT-Immediate, RAVLT-Learning, RAVLT-Forgetting, and RAVLT-Percent Forgetting). For each group, we first calculated its average scores on each of the cognitive measures, and since the tests use different metrics, we then scaled and centered the values across all the groups. We used the Complex Heatmap package’s default clustering method (distance metric = ‘Euclidean’, linkage method = ‘complete’) ^30^ to measure the proximity between subtypes along with the progression from CN to AD, based on their cognitive measures.

### 2.8 Trajectory analysis for ADNI patient subgroups using clinical information

Trajectory analysis represents an ordering of patient subtypes based on consecutive states of disease progression, from a completely normal state (root node) to a final state (end node). For any clinical trajectory, pseudo-time reconstruction quantifies the progression between states or nodes based on the data used in the analysis. In this study, to evaluate the severity and order of dementia progression, we compared the cognitive scores of EMCI and LMCI subtypes with those of CN and AD patients in the ADNI cohort. Then we used the ‘ClinTrajan’ python package ^31^ to quantify the pseudo time of dementia progression states based on the cognitive measures. We set the parameters for principal tree calculation as several nodes = 40, α=0.01, μ=0.1, λ=0.05. We chose the trajectory that goes from CN patients (with the least dementia risk) as root nodes to AD patients (highest level dementia and cognitive impairment).

### 2.9 Time-to-event (dementia diagnosis) analysis in MCI patients of ADNI dataset

Time-to-event survival analysis shows the differences in the probabilities of dementia between each state during the progression of the disease. We used the longitudinal clinical diagnosis data from ADNI with three labels: ‘Control’, ‘MCI’ and ‘Dementia’ and checked the latest diagnosis of each patient after their first visit every six months. Then we used their latest diagnosis as the event at each checkpoint and calculated the percentage of the label ‘Dementia’ among all of the three labels. At last, we plotted the dementia percentage against each checkpoint time for 96 months.

### 2.10 Scaled Gene Score with Common Differential Expressed Genes

Scaled gene-set score indicates the collective activities of genes within a group. 25 genes are commonly up-regulated and 6 genes are commonly down-regulated, between the EMCI-1 vs EMCI-2 DE analysis and LMCI-1 vs LMCI-2 DE analysis in the ADNI dataset. These genes are selected for entropy-based gene set scoring, as proposed previously ^32^. A t-test was performed between the gene scores of the EMCI-1 vs. EMCI-2 and LMCI-1 vs. LMCI-2 subgroups. The formulas for calculating the entropy estimation and scaled gene scores are shown below:

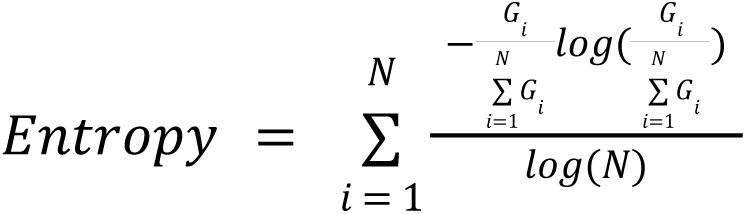

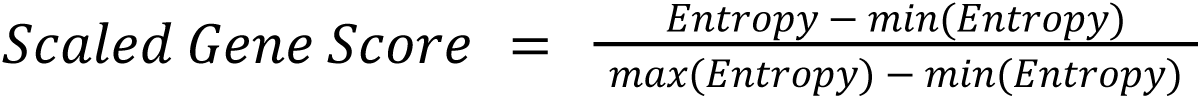

### 2.11 EFIGA dataset for validation

The Estudio Familiar de Influencia Genetica en Alzheimer (EFIGA) is a study recruiting individuals between January 1, 2018, and April 30, 2022, with suspected sporadic and familial AD and healthy controls similar in age, in the Dominican Republic and in the Washington Heights neighborhood of New York City ^33^. The diagnosis of Alzheimer’s clinical syndrome was made according to NIA-AA criteria ^34^. All clinical diagnoses were established through a consensus conference involving a neurologist, a neuropsychologist, and an internist specializing in dementia and geriatrics. Patients’ blood was collected and plasma metabolomics were measured by Thermo Orbitrap HFX Q-Exactive mass spectrometer. Metabolomics data were processed through a computational pipeline that leverages open source feature detection and peak alignment software, apLCMS52 and xMSanalyzer ^35,36^.

### 2.12 Label transferring validation of ADNI dataset by EFIGA dataset

We verified the robustness of EMCI vs LMCI inference, EMCI -1 vs EMCI-2 and LMCI-1 vs LMCI-2 patient subtype inferences, by the EFIGA dataset from Columbia University^37^. We first selected common metabolites (n=54) between the two cohorts per HMDB ID based matching. Before label transferring, we ensured data quality and consistency by applying a log₂ transformation, removing outliers, scaling the data to center each metabolite’s distribution, and clipping values beyond ±3 standard deviations. We also filtered out metabolites with high variability (CV > 0.2), low reproducibility (ICC < 0.65), and low confidence (confidence level = 5) in the EFIGA cohort, and consolidated biological replicates by averaging their values. For label transfering, we used Unsupervised Topological Alignment for Single-Cell Multi-Omics Integration (UnionCom) ^38^, which aligns datasets by leveraging an adjacency matrix to assess inter-data similarities (distance mode = default ‘geodesic distance’ for EMCI and LMCI transferring, distance mode = ‘l2’ for further subtypes) ^39^. After normalizing both datasets using MinMax separating, we applied Unioncom separately for EMCI and LMCI—first inferring EMCI1 vs. EMCI2 and LMCI1 vs. LMCI2—and then combining these to obtain the final patient subtype labels for best sensitivity. A 99.5% similarity threshold was implemented to match the patient labels between the two cohorts and recovered the most similar patient sample in EFIGA data. For the inferred patient labels in the EFIGA data set, we tested possible differences in the plasma biomarkers Aβ42, Aβ40, p-Tau181 and p-Tau217 ^37^, using the Wilcoxon test and chi-squared test. In addition to plasma biomarkers, p-Tau181 levels were dichotomized at the cutoff point of 2.63 pg/mL, based on the previous study, to classify patients as Alzheimer’s disease-positive or negative ^37^.

## 3. RESULTS

### 3.1 Mutliomics integration through similarity network fusion

First, considering there are many more transcriptomics features than metabolomics features, we reduced the dimensionality of the transcriptomics dataset from 20,032 to 172 using a stacked autoencoder and set the bottleneck layer size to 172 (**Figure 1A**). Then transcriptomics data were split into train and test sets in a ratio of 80:20. We trained the autoencoder on the training set and evaluated the Mean Squared Error (MSE) metric on the test set (Test Set - MSE: 0.1175, suggesting a high fidelity representation of the transcriptomics data. Our model training procedure minimized the MSE metric at epoch 10 (**Figure 1B)**, Hence we saved the model weights at epoch 10 for further use. For data integration, we extracted the output of the autoencoder bottleneck layers, which represented a low-dimensional representation of the transcriptomics data. We calculated the patient affinity matrix for both transcriptomics and metabolomics as the input to the similarity network fusion method (**Figure 1C**) ^22^.

**Figure 1.**
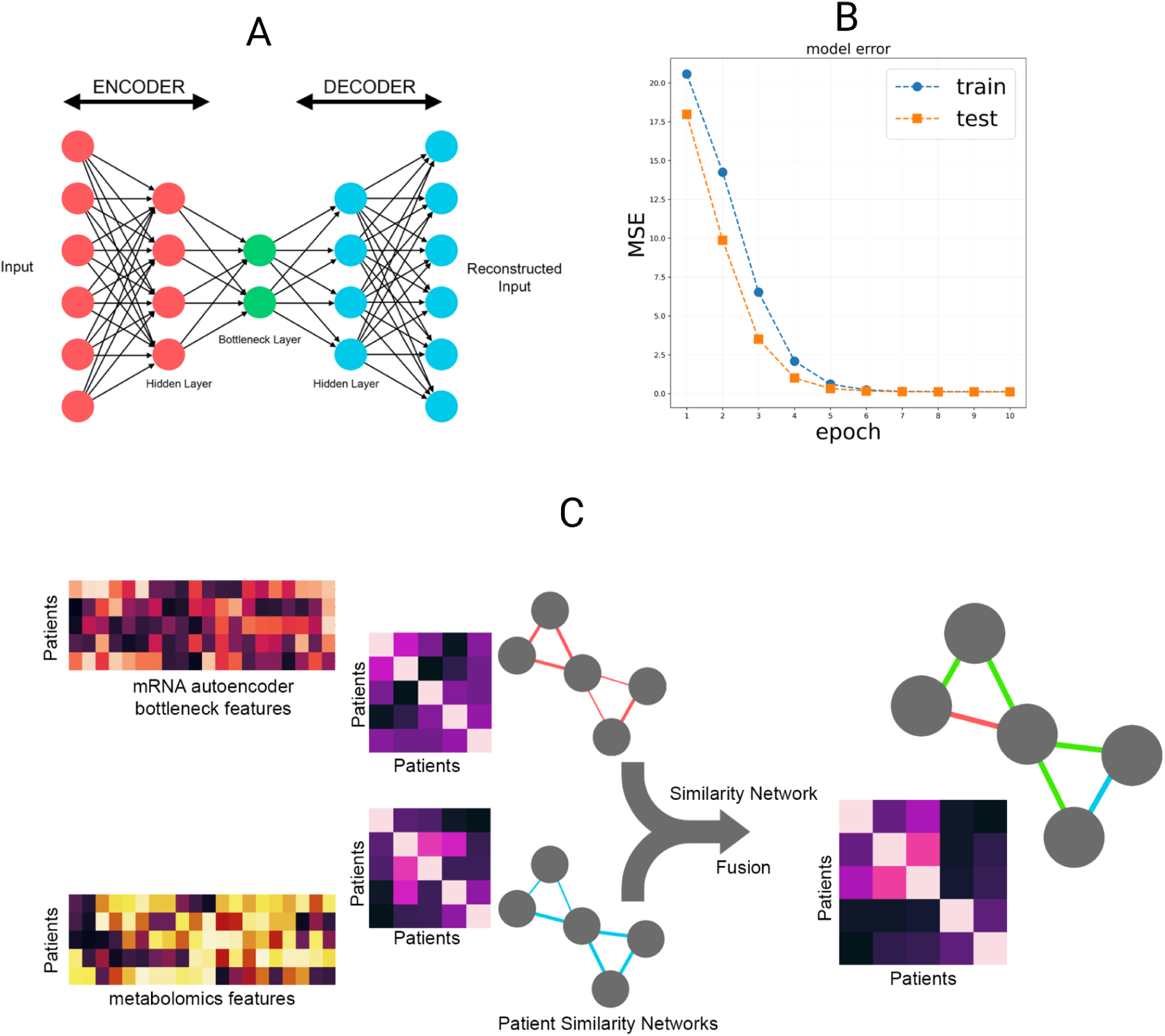
Procedures for integrating metabolomics data with transcriptomics data. (A) Dimensionality reduction for transcriptomics data, using stacked autoencoders. The number of bottleneck layers of features are designed to be the same as the metabolomics features. (B) Mean squared error for both training and test set vs training epoch for the stacked autoencoder showing that the model has converged. (C) Integrating transcriptomics and metabolomics with similarity network fusion (SNF), which are then used to identify subtypes in EMCI and LMCI, respectivel

Next, we performed spectral clustering on the SNF-integrated patient affinity matrix in the EMCI cohort. We varied the number of clusters from 2 to 10 and calculated the SC and CH scores, two metrics measuring the fitness of the subclusters, respectively. We found that in the multi-omics case, the optimum number of clusters is 2 for both metrics (**Figure 2A-B**). We also tested other clustering methods besides spectral clustering, such as K-mean, hierarchical clustering and Gaussian mixed model clustering, and found that spectral clustering gave the best subgroup separation **(Supplementary Figure 1**). We investigated the contribution of the gene expression vs. metabolomics on integrated subtypes by Silhouette scores. Metabolomics and gene expression has Silhouette score of 0.41 and 0.21, indicating that metabolomics data are the major contributor to the subtypes after SNF **(Supplementary Figure 2).**

**Figure 2.**
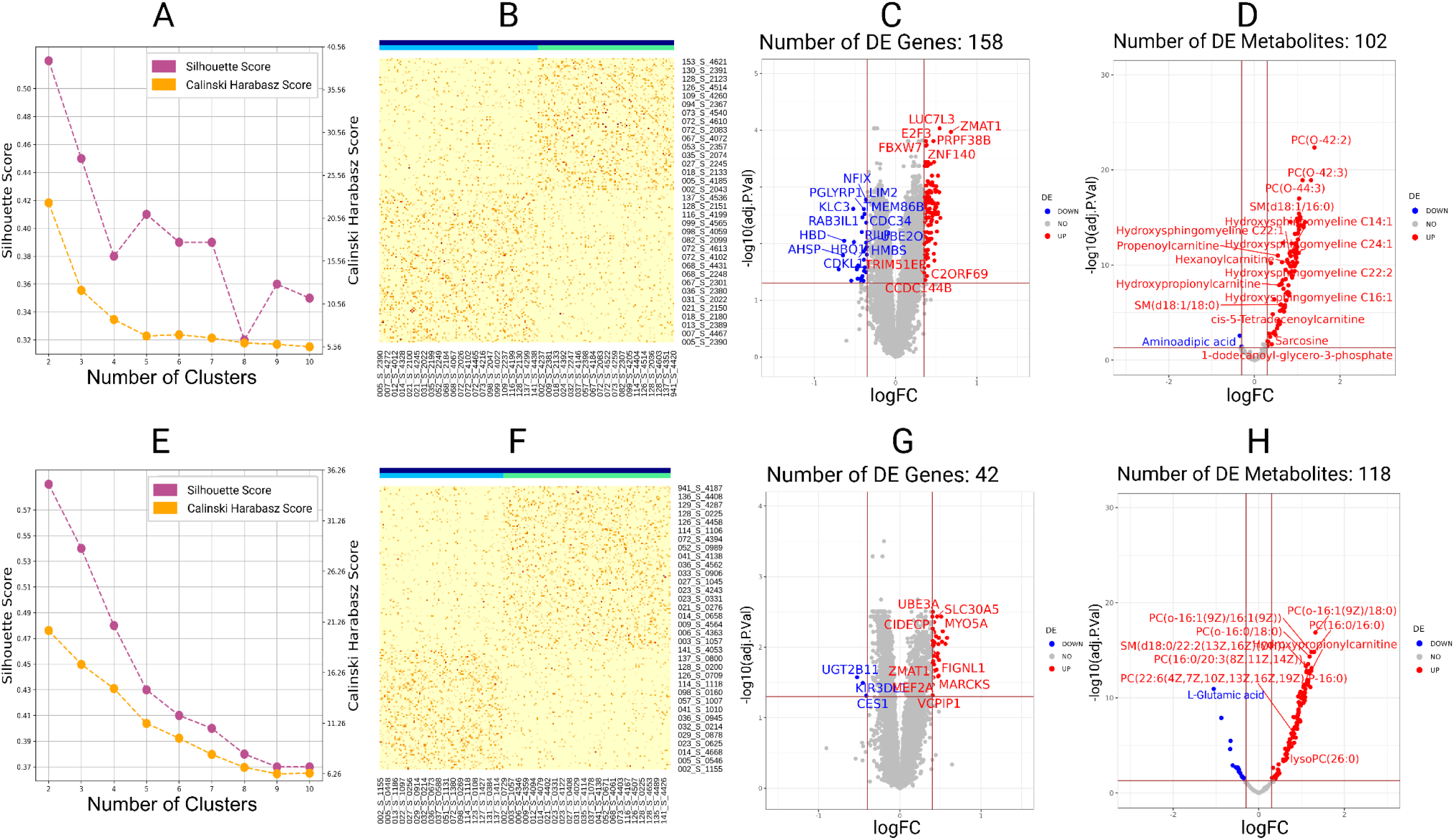
Multi-omics analyses of the **EMCI** group (top) and the **LMCI** group (bottom) (A) Estimation of optimum number of clusters (from spectral clustering) in SNF integrated data in EMCI samples. Silhouette score and Calinski Harabasz score are used as metrics. (B) Visualization of patient-level spectral clustering results for the two clusters based on SNF-integrated data. (C-D) Differentially expressed genes (C) and differential metabolites (D) for the two clusters, with the less severe subtype EMCI-1 as the baseline control. (E-H) plots are done the same way as in (A-D), but for the LMCI group, where the less severe LMCI-1 is set as the baseline control.

To characterize the marker genes and metabolites that differentiate the subtypes, we performed differential expression (DE) gene analysis and DE metabolite analysis **(Figure 2C, 2D**). For DE gene analysis, we used limma with adjustment on the blood cell types and clinical covariates such as age, gender and race. For the DE plasma metabolite analysis, we adjusted by clinical covariates data. We named the less severe subtype EMCI-1 as the baseline (control), relative to EMCI-2 subtype (case). The top ten upregulated DE genes are *LUC7L3, ZMAT1, PRPF38B, E2F3, ZNF140, FBXW7, KRCC1, ZNF420, TBK1, ZNF146* **(Table 2**; **Figure 2C).** The top ten differentially expressed metabolites are primarily phosphatidylcholine and sphingomyelins (**Table 2**; **Figure 2D)**.

**Table 2.** Top 10 Differentially Expressed Genes and Metabolites in Early Mild Cognitive Impairment (EMCI) and Late Mild Cognitive Impairment (LMCI) Groups ( (Absolute Fold Change>1.2 for genes, >1.3 for metabolites, P value < .001).

|  | DE Genes | DE Metabolites |
| --- | --- | --- |
| **Early Mild Cognitive Impairment** | LUC7L3<br>ZMAT1<br>PRPF38B<br>E2F3<br>ZNF140<br>FBXW7<br>KRCC1<br>ZNF420<br>TBK1<br>ZNF146 | PC(o-16:1(9Z)/18:0)<br>PC(16:0/16:0)<br>PC(o-16:1(9Z)/16:1(9Z))<br>PC(o-16:0/18:0)<br>PC(o-18:0/18:2(9Z,12Z))<br>SM(d18:0/22:2(13Z,16Z)(OH))<br>Butenylcarnitine<br>Hydroxypropionylcarnitine<br>PC(16:0/20:3(8Z,11Z,14Z))<br>SM(d18:0/14:1(9Z)(OH)) |
| **Late Mild Cognitive Impairment** | UBE3A<br>MYO5A<br>SLC30A5<br>CIDECP<br>ZNF23<br>SELT<br>MYBL1<br>GLCCI1<br>ADO<br>PDIK1L | PC(O-42:2)<br>PC(O-42:3)<br>PC(O-44:3)<br>SM(d18:1/16:0)<br>PC(O-34:1)<br>PC(O-18:1(9Z)/18:2(9Z,12Z))<br>PC(36:1)<br>PC(O-36:1)<br>PC(18:3(6Z,9Z,12Z)/22:0)<br>PC(O-18:0/18:0) |

Similarly to EMCI, we performed spectral clustering on the SNF-integrated patient similarity matrix in the LMCI group (**Figure 2E, F**). Using the less severe subtype LMCI-1 as the control, we identified 42 DE genes and 118 DE metabolites. (**Figure 2G, H**). Among them, 19 genes overlap with the DE genes in EMCI, and 84 metabolites overlap with the DE metabolites in EMCI. The top ten differentially expressed genes between the two LMCI subpopulations are *UBE3A, MYO5A, SLC30A5, CIDECP, ZNF23, SELT, MYBL1, GLCCI1, ADO, PDIK1L* (**Table 2**;**Figure 2G**). The top ten differentially expressed metabolites are phosphatidylcholine and sphingomyelins too (**Table 2**; **Figure 2H**). The highly similar DE metabolites between LMCI and EMCI are consistent with the expectation that metabolomics are the main player in SNF based subtyping (**Supplementary Figure 3**).

### 3.2 Gene set and metabolomic enrichment analysis of EMCI subgroups

We next conducted GSEA analysis of the DE genes in the EMCI group, and found 5 activated and 5 suppressed Gene Ontology (GO) terms (*P* value<.05). The activated GO terms are DNA repair, positive regulation of multicellular organismal process, cytoplasmic vesicle, intracellular vesicle and biological process involved in interspecies interaction between organisms (**Figure 3A**). The suppressed GO terms are myeloid cell differentiation, plasma membrane bounded cell projection organization, cell projection organization, cell projection assembly, and plasma membrane bounded cell projection assembly (**Figure 3A**). Such patterns indicate that the vesicle transportation is active whereas the blood cells are rounding up. The bipartite graph of activated or suppressed pathways and their leading edge genes for the EMCI subtypes is shown in **Figure 3B**. *PLEK2, RAP1GAP, KLC3*, *CDKL1, ESPN,* and *RILP* are the genes shared by the suppressed pathways associated with cell projection. *PALLD* and *TSPAN2* are shared by the activated pathways.

**Figure 3.**
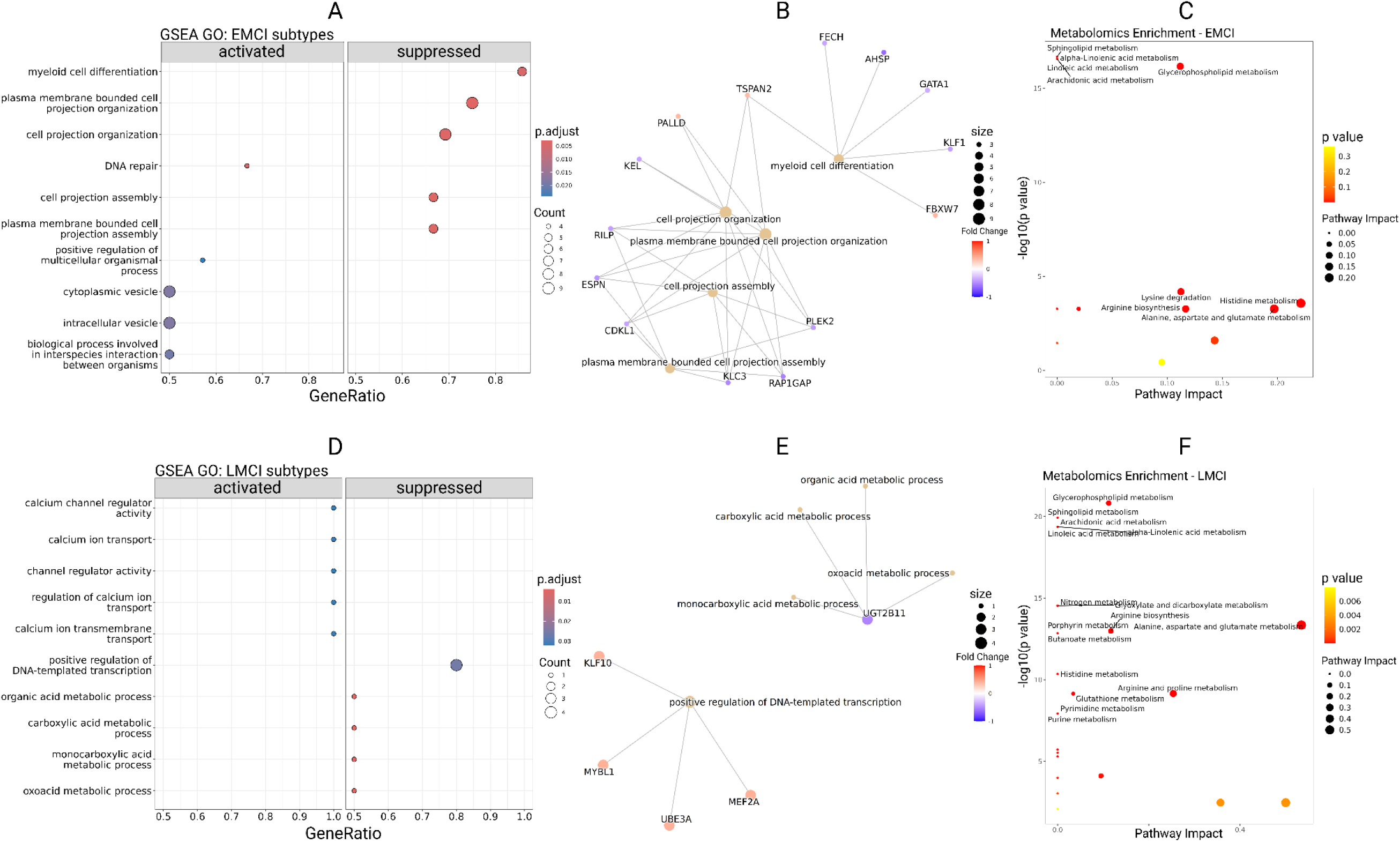
Gene set enrichment analysis (GSEA) of activated and suppressed pathways between **EMCI-1** vs. **EMCI-2** (top) and **LMCI-1** vs. **LMCI-2** (bottom) using the differentially expressed (DE) genes between the two subtypes. (A) Dysregulated pathways based on DE genes in the EMCI subtypes, where EMCI-1 is the baseline condition. (B) Bipartite results show the DE genes associated with the dysregulated pathways in the EMCI subtypes. (C) Enriched metabolic pathways between the EMCI subtypes. (D-F) The same analyses as done in (A-C), but in LMCI subtypes, with LMCI-1 set as the baseline condition.

For metabolomics pathways, EMCI-2 shows 2 activated and 5 suppressed pathways compared to EMCI-1 (**Table 3**; **Figure 3C**). The activated pathways are glycerophospholipid metabolism and tryptophan metabolism. The suppressed pathways are lysine degradation, histidine metabolism, glutathione metabolism, alanine, aspartate and glutamate metabolism, and arginine biosynthesis. L-glutamate, phosphatidylcholine and L-tryptophan are metabolites with the most connections with these pathways.

**Table 3.**
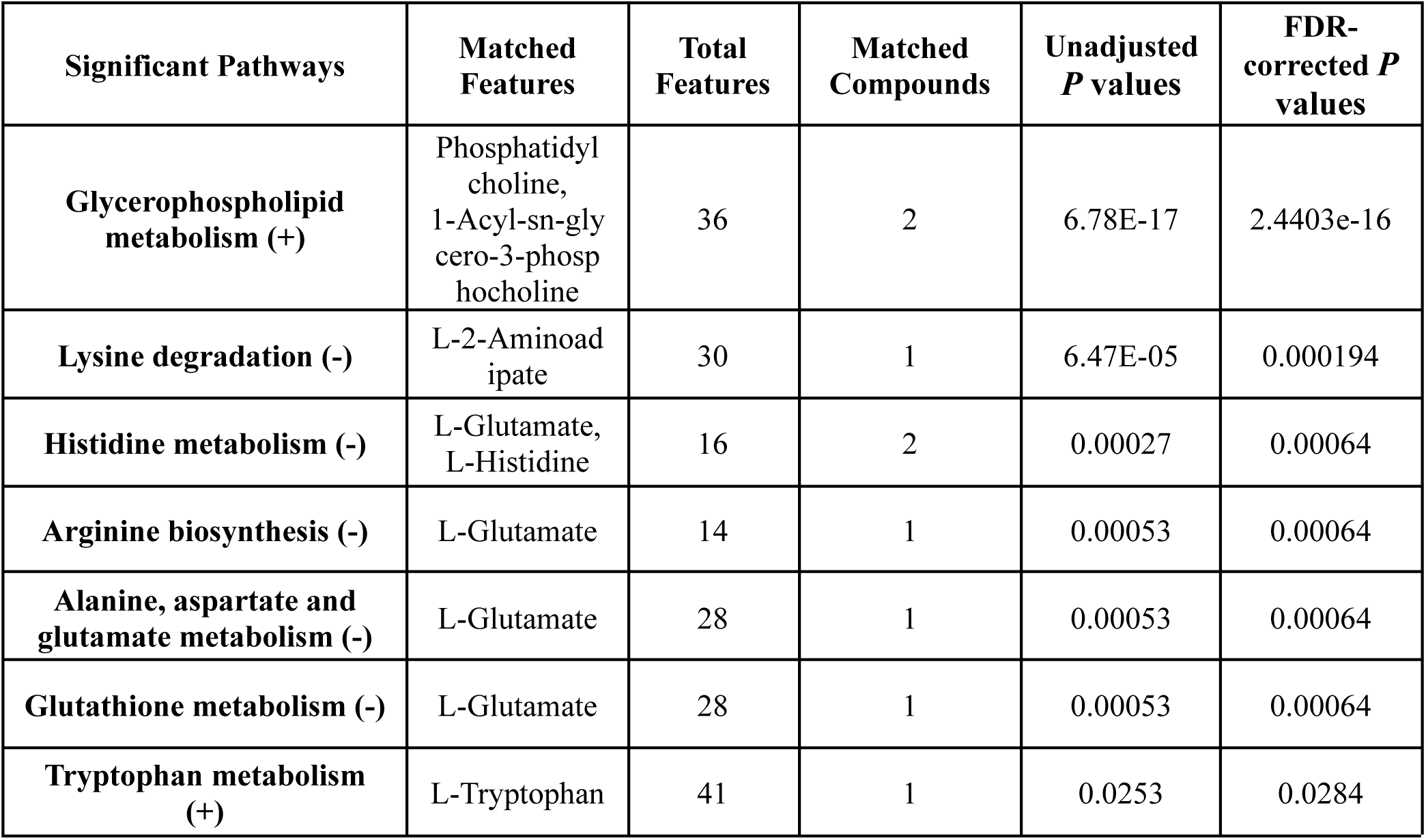
Significantly activated (+) or suppressed (-) metabolomic pathways in the EMCI-2 subtype compared to EMCI-1, based on metabolite enrichment analysis.

| **Significant Pathways** | **Matched Features** | **Total Features** | **Matched Compounds** | **Unadjusted *P* values** | **FDR-corrected *P* values** |
| --- | --- | --- | --- | --- | --- |
| **Glycerophospholipid metabolism (+)** | Phosphatidyl choline, 1-Acyl-sn-glycero-3-phosphocholine | 36 | 2 | 6.78E-17 | 2.4403e-16 |
| **Lysine degradation (-)** | L-2-Aminoadipate | 30 | 1 | 6.47E-05 | 0.000194 |
| **Histidine metabolism (-)** | L-Glutamate, L-Histidine | 16 | 2 | 0.00027 | 0.00064 |
| **Arginine biosynthesis (-)** | L-Glutamate | 14 | 1 | 0.00053 | 0.00064 |
| **Alanine, aspartate and glutamate metabolism (-)** | L-Glutamate | 28 | 1 | 0.00053 | 0.00064 |
| **Glutathione metabolism (-)** | L-Glutamate | 28 | 1 | 0.00053 | 0.00064 |
| **Tryptophan metabolism (+)** | L-Tryptophan | 41 | 1 | 0.0253 | 0.0284 |

### 3.3 Gene set and metabolomic enrichment for the LMCI subtypes

We performed similar systems analysis of the genes and metabolites in the LMCI-2 vs LMCI-1 subtypes. The GESA 5 significantly activated GO terms and 5 significantly suppressed GO terms were found (**Figure 3D**). The 5 activated pathways are calcium channel regulator activity, calcium ion transport, channel regulator activity, regulation of calcium ion transport, and calcium ion transmembrane transport. The 5 suppressed pathways are positive regulation of DNA-templated transcription, organic acid metabolic process, carboxylic acid metabolomic process, monocarboxylic acid metabolomic process, and oxoacid metabolic process. Such patterns indicate that the calcium channel is active whereas the carboxylic acid metabolomics is repressed, very different from the emerging functional changes in EMCI subtypes. *KLF10, MEF2A, MYBL1, UBE3A* are the key genes associated with the 5 activated pathways (**Figure 3E**). On metabolomics data, we detected pathways 2 activated and 6 suppressed pathways in LMCI-2 compared to LMCI-1 (**Table 4**; **Figure 3F**). The activated pathways are glycerophospholipid metabolism, and glutathione metabolism. The suppressed pathways are alanine, aspartate and glutamate metabolism, arginine biosynthesis, arginine and proline metabolism, glycine serine and threonine metabolism, phenylalanine tyrosine and tryptophan biosynthesis, and phenylalanine metabolism. L-glutamate, L-aspartate, and L-phenylalanine are metabolites with the most connections with pathways in **Figure 3F**.

**Table 4.** Significantly activated (+) metabolomic pathways in the LMCI-2 subtype, compared to LMCI-1, based on metabolite enrichment analysis.

| **Significant Pathways** | **Matched Features** | **Total Features** | **Matched Compounds** | **Unadjusted *P* values** | **FDR-corrected *P* values** |
| --- | --- | --- | --- | --- | --- |
| **Glycerophospholipid metabolism (+)** | Phosphatidylcholine, 1-Acyl-sn-glycero-3-phosphocholine | 36 | 2 | 1.51E-21 | 3.93E-20 |
| **Alanine, aspartate and glutamate metabolism (-)** | L-Aspartate, L-Alanine, L-Glutamate, L-Glutamine | 28 | 4 | 4.48E-14 | 1.46E-13 |
| **Arginine biosynthesis (-)** | L-Glutamate, L-Aspartate, L-Glutamine | 14 | 3 | 1.02E-13 | 1.84E-12 |
| **Arginine and proline metabolism (-)** | L-Glutamate, Putrescine, Spermidine | 36 | 3 | 7.03E-10 | 1.3E-09 |
| **Glutathione metabolism (-)** | L-Glutamate, Putrescine, Spermidine | 28 | 3 | 7.03E-10 | 1.3E-09 |
| **Glycine, serine and threonine metabolism (-)** | Sarcosine, L-Threonine | 33 | 2 | 7.63E-05 | 9.93E-05 |
| **Phenylalanine, tyrosine and tryptophan biosynthesis(-)** | L-Phenylalanine | 4 | 1 | 0.00338 | 0.00351 |
| **Phenylalanine metabolism (-)** | L-Phenylalanine | 8 | 1 | 0.00338 | 0.00351 |

### 3.4 Validation of new EMCI and LMCI subtypes by ADNI clinical phenotypes

We first compared the cognitive measures among the subtypes and found a clear progression from healthy controls to AD (**Figure 4A**). For some cognitive measures, a lower score means cognitively healthier such as ADAS-11, ADAS-13, ADAS-Q4, RAVLT-Forgetting, RAVLT-Percent Forgetting, CDRSB, FAQ, TRABSCOR). For other measures, a higher score means cognitively healthier, such as RAVLT-Learning, RAVLT-Immediate, LDELTOTAL, and MMSE. As shown in the heatmap clustering, there is a clear transition from the controls, through EMCI-1, EMCI-2 and LMCI-1, LMCI-2 to AD patients. Control patients show the lowest scores for ADAS-11, ADAS-13, ADAS-Q4, RAVLT-Forgetting, RAVLT-Percent Forgetting, CDRSB, FAQ, and TRABSCORE, and the highest scores for RAVLT-Learning, RAVLT-Immediate, LDELTOTAL, and MMSE. Progressing from healthy controls to EMCI-1, EMCI-2, LMCI-1, LMCI-2, and AD, this pattern gradually reverses (**Table 5**). Interestingly, the EMCI-2 and LMCI-1 subtypes are clustered next to one another, further demonstrating the fine-grained subtle difference between EMCI and LMCI subtypes, in the continuum of AD disease progression.

**Figure 4.**
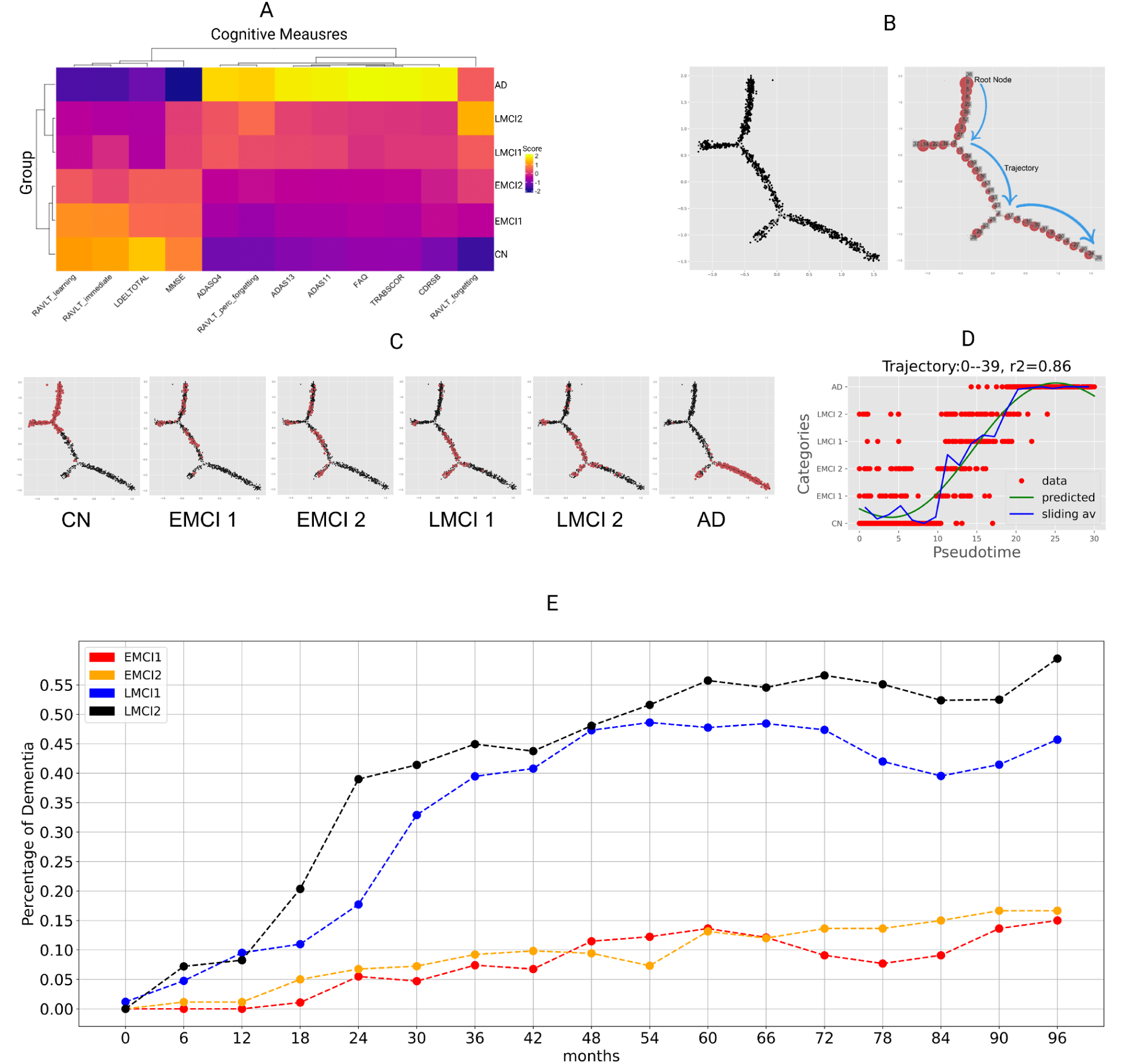
Validation of the subtypes by clinical and phenotypic information. (A) Hierarchical clustering-based comparison of the EMCI and LMCI subtypes with ‘Control’ and ‘Dementia’ patients based on cognitive measures. (B) Principal tree illustrating the multidimensional structure in the Alzheimer’s dataset. Visualization of disease progression trajectory.. Controls are shown as the root node and the patients with the highest risk of dementia are shown as the final state. (C) Ordered patient phenotypes from ‘Control’, EMCI-1, EMCI-2, LMCI-1, LMCI-2 and AD(dementia) on the trajectory tree. The consecutive states of progression of Alzheimer’s disease are evident. (D) Clinical trajectory regression analysis for Trajectory 0-39 (Control-MCI-Dementia) with pseudotime representing the degree of disease progression along the trajectory. (E) Time-to-diagnosis plot for the percentage of dementia in every six-month check-up, among the EMCI and LMCI subtypes.

**Table 5.** Scores for each group on cognitive measures. Cognitive measures include the Clinical Dementia Rating (CDR), the Alzheimer’s Disease Assessment Scale (ADAS-11, ADAS-13, ADAS-Q4), the Mini Mental State Exam (MMSE), subtests from the Rey Auditory Verbal Learning Test (RAVLT-Immediate, RAVLT-Learning, RAVLT-Forgetting, RAVLT-Percent Forgetting), the Delayed Recall Total score (LDELTOTAL), the Trail Making Test (TRABSCOR), and the Functional Activities Questionnaire (FAQ). Additional demographic and clinical information includes the total number of cases for each stage (Total Cases), the number of females in each stage (Female), and the mean age of participants (Age).

|  | CDRSB | ADAS11 | ADAS13 | ADASQ4 | MMSE | RAVLT<br>Immediate | RAVLT<br>Learning | RAVLT<br>Forgetting | RAVLT<br>Percent<br>Forgetting | LDELTOT<br>AL | TRABSCO<br>R | FAQ | Total<br>Cases | Femal<br>e | Age |
| --- | --- | --- | --- | --- | --- | --- | --- | --- | --- | --- | --- | --- | --- | --- | --- |
| CN | 0.0318 | 5.8769 | 9.0548 | 2.7640 | 29.0581 | 45.3590 | 5.9436 | 3.5789 | 33.7744 | 13.2828 | 82.3616 | 0.1248 | 534 | 158 | 73.454<br>4 |
| EMCI-1 | 1.1505 | 7.4782 | 11.7826 | 3.8387 | 28.4516 | 43.3548 | 5.7634 | 4.1183 | 41.0667 | 9.1613 | 92.75 | 1.5435 | 108 | 32 | 72.052<br>8 |
| EMCI-2 | 1.3101 | 8.3611 | 13.3704 | 4.3704 | 28.1944 | 36.9167 | 4.7870 | 4.3426 | 49.8148 | 8.6204 | 101.2 | 2.1574 | 93 | 62 | 69.448<br>4 |
| LMCI-1 | 1.7 | 11.1247 | 17.7595 | 5.9739 | 27.5391 | 34.4783 | 3.6348 | 4.4870 | 61.3610 | 4.1913 | 117.7544 | 3.6957 | 85 | 8 | 75.109<br>4 |
| LMCI-2 | 1.5706 | 10.6551 | 17.3454 | 5.8706 | 27.4353 | 30.8471 | 3.4824 | 4.8824 | 68.7140 | 4.1294 | 116.2143 | 4.0235 | 115 | 68 | 72.379<br>1 |
| AD | 4.4248 | 19.6361 | 29.9617 | 8.6283 | 23.1947 | 22.8244 | 1.8422 | 4.4940 | 89.1390 | 1.3392 | 197.4188 | 13.224<br>8 | 339 | 152 | 74.987<br>6 |

In order to validate the progression of severity among the subtypes, we computed the clinical trajectory using the elastic principal tree (EPT) algorithm. For this, we concatenated the cognitive measures from the EMCI and LMCI subtypes with those from dementia patients and healthy controls. The principal tree comprises an assembly of principal curves representing the topology between the samples ^40^. We set the trajectory root node as the healthy control patients with the least risk of dementia (**Figure 4B**). EPT allows us to visualize the location of patients in each category on the tree, from healthy control to MCI and AD. There is a gradual shift in patients’ risk categories while traversing along the trajectory, illustrating the consecutive states of dementia progression among the subtypes (**Figure 4C**). There is also a high correlation (r2 value = 0.86) between the dementia subtypes along the categories and the reconstructed pseudotime (**Figure 4D**). These results again validated the gradual shift in dementia risk in the order of the patients from CN, through EMCI-1, EMCI-2, LMCI-1, LMCI-2 to AD, as reflected by the trajectory of dementia progression.

Lastly, to associate the new subtypes with AD disease risks, we also examined the percentage of the patients in each subtype who were later clinically diagnosed as having “dementia” in the following check-ups every six months since their first screening (**Figure 4E**). Again there is a trend of increasing percentages of dementia in each check-up time, when the states change from the mildest EMCI-1 stage to the most severe stage of LMCI-2. Thus, different analyses confirmed the clinical relevance of the four subtypes within the MCI subjects in ADNI.

### 3.5 Validation of EMCI and LMCI with EFIGA dataset

We validated our EMCI vs. LMCI patient inference using the EFIGA dataset (N = 717) from Columbia University ^37^ (**Figure 5**). We built a UnionCom-based label-transferring model ^38^ using the 54 overlapping metabolites between ADNI and EFIGA datasets. We inferred 536 EMCI patients and 181 LMCI patients from the EFIGA cohort. Differential expression analysis identified 18 significantly altered metabolites in EFIGA datasets (**Figure 5A**). Among these, 14 metabolites overlapped with the differential metabolites in ADNI data, with 8 showing consistent directions of changes (**Figure 5B**). These include 3-Hydroxy-cis-5-tetradecenoylcarnitine, Tryptophan, 3-Hydroxy-5,8-tetradecadien oyl carnitine, Decanoylcarnitine, Stearoylcarnitine, Lauroyl Carnitine, PC(40:5), and Octanoylcarnitine. The UMAP plot shows distinct patterns between EMCI and LMCI groups in EFIGA data (**Figure 5C**). To validate the inferred EMCI and LMCI patient labels, we compared the biomarkers levels between the two groups. p-Tau181 (phosphorylated tau at threonine 181) is a biomarker associated with AD, contributing to neurofibrillary tangles, a hallmark of AD. p-Tau181 levels are significantly higher in the LMCI group, compared to EMCI (**Figure 5D**). Furthermore, other protein markers, including p-Tau217, Aβ42, and Aβ40, also show significantly higher levels in the LMCI group compared to the EMCI group, with p-Tau181 showing particularly marked elevation (*P < 0.05*) (**Figure 5E-H**).

**Figure 5.**
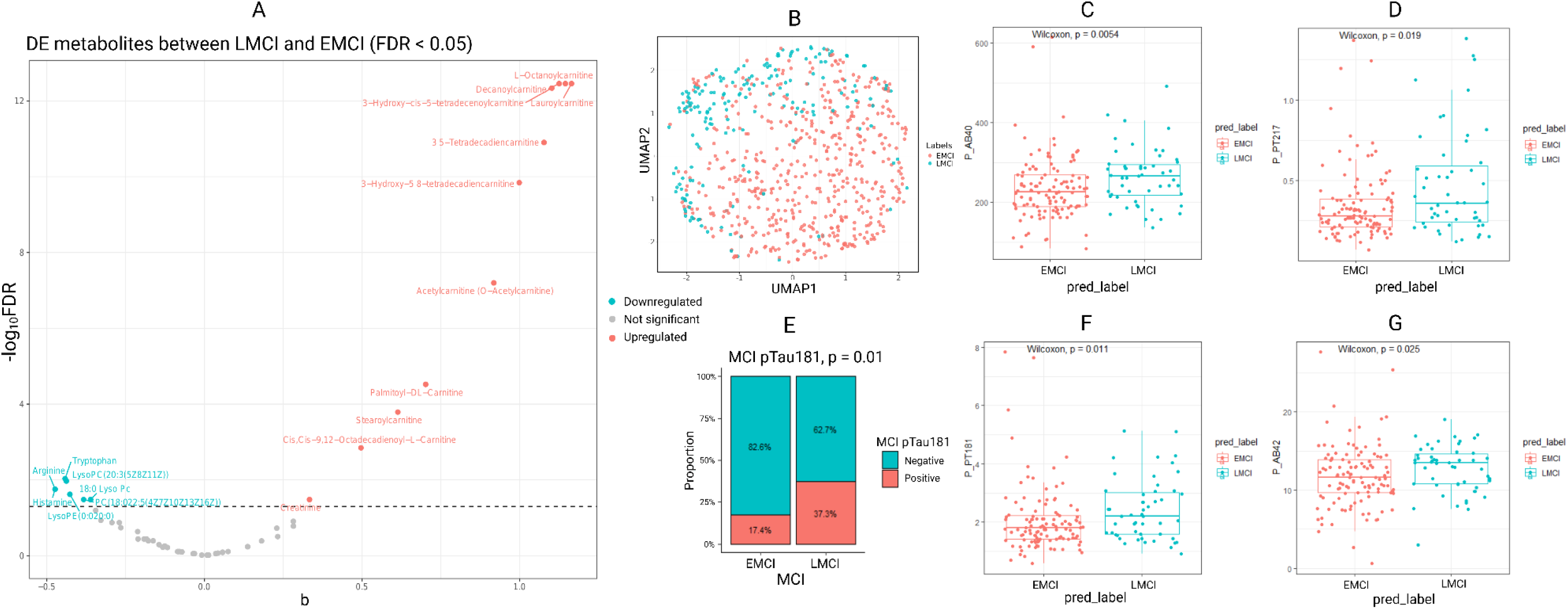
Validation of EMCI vs LMCI classification by metabolomics data from EFIGA cohort. (A) Differentially expressed metabolites between EMCI and LMCI. (B) Comparison of the differential metabolites between EFIGA and ADNI cohorts. (C) UMAP plot comparing EMCI vs LMCI. (C) p-Tau181 binary status for MCI patients. (E) Aβ40 EMCI vs LMCI boxplot. (F) p-Tau217 EMCI vs LMCI boxplot. (G) p-Tau181 EMCI vs LMCI boxplot. (G)Aβ42 EMCI vs LMCI boxplot.

### 3.6 Validation of new EMCI and LMCI subtypes with EFIGA dataset

We further examined whether the EMCI-1/2 and LMCI-1/2 subtypes identified in ADNI could be validated in the EFIGA dataset. We applied Unioncom subtype labels transferred from ADNI to the above inferred EMCI and LMCI samples in EFIGA. In total we identified 176 EMCI-1, 360 EMCI -2, 116 LMCI-1, and 65 LMCI-2 patients respectively. Visualization of the subtypes through UMAP shows clear separation patterns between the two LMCI subtypes, while EMCI subtypes have more subtle distinctions as expected **(Figure 6A)**. Impressively, p-Tau181 positivity increases progressively among the four subtypes, with LMCI 2 exhibiting the highest proportion of p-Tau181 positive samples (50%) compared to EMCI-1 (15.8%) which has the lowest presence of p-Tau181 positivity (**Figure 6B**). This aligns with expected pathological progression ^41^.

**Figure 6.**
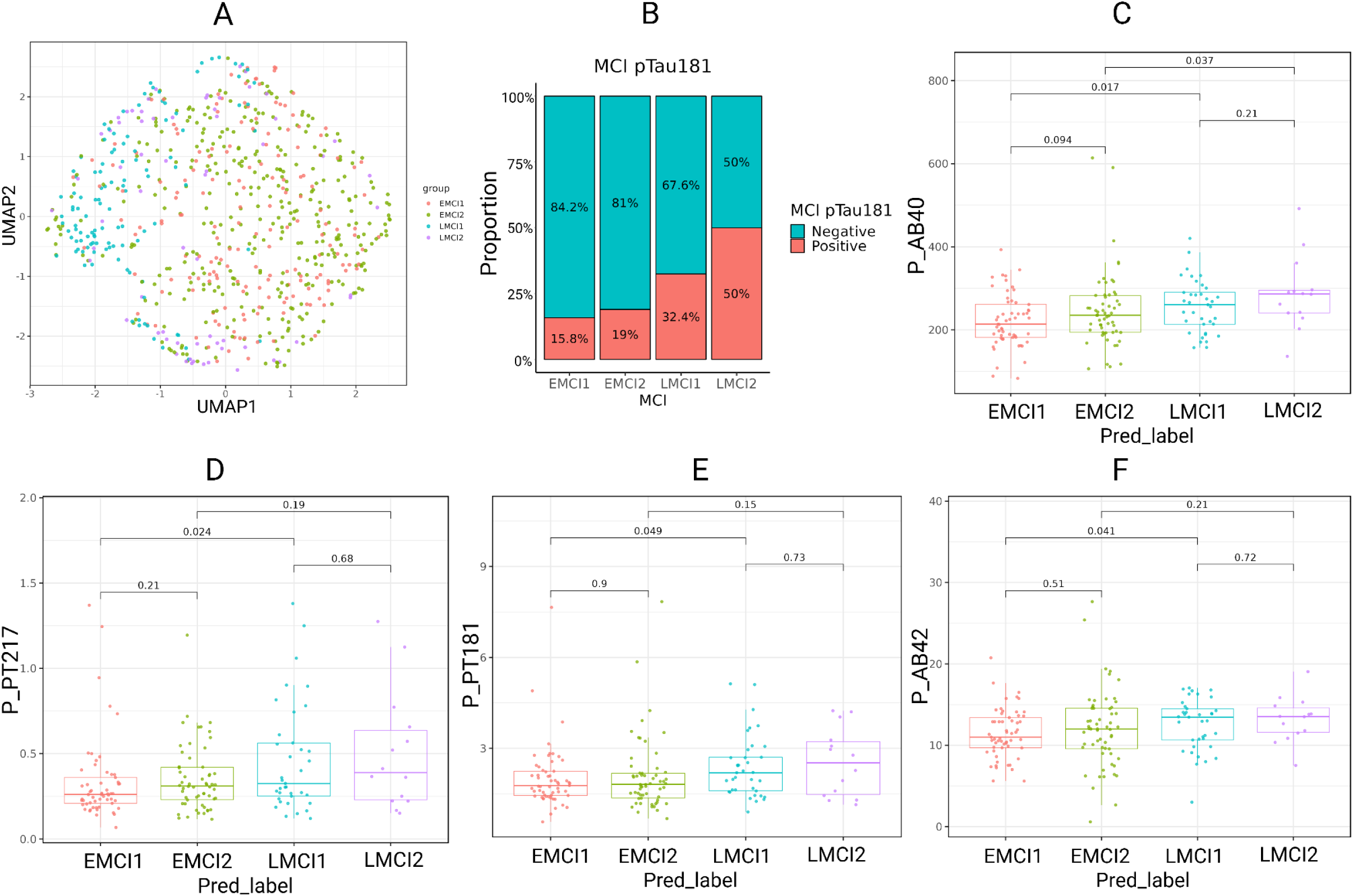
Validation of the further subtypes by metabolomics data from EFIGA. (A) UMAP plot comparing the four 4 MCI subtypes. (B) p-Tau181 Status for MCI refined subtypes patients. (C) Aβ40 EMCI1 vs EMCI2 vs LMCI1 vs LMCI2 boxplot. (D) p-Tau217 EMCI1 vs EMCI2 vs LMCI1 vs LMCI2 boxplot. (E) p-Tau181 EMCI1 vs EMCI2 vs LMCI1 vs LMCI2 boxplot. (F)Aβ42EMCI1 vs EMCI2 vs LMCI1 vs LMCI2 boxplot.

Further analyses of the other biomarkers in the refined subtypes also reveal consistent biomarker patterns across groups, with a notable progressive trend from early to late stages. Aβ40, p-Tau217 p-Tau181, and Aβ42 levels are significantly different between EMCI-1 and LMCI-1 (**Figure 6C-F**). Additionally, Aβ40 also shows significant differences between EMCI-2 vs LMCI-2 *(P < 0.05)* and almost significant difference between EMCI-1 and EMCI-2 **(Figure 6C)** ^42^. This suggests that while the above measured biomarkers maintain stability within early or late subtypes, Aβ40 may serve as a more sensitive indicator of molecular changes between subgroups ^43^.

## 4. DISCUSSION

The current study aims to identify clinically relevant molecular subtypes in the MCI cohort of AD patients, by integrating metabolomics and gene expression data. We focused on patients in the MCI stage, as it is a sensitive window important for therapeutic intervention. We found that both EMCI and LMCI can be further split into two relatively even subtypes, which we call EMCI-1, EMCI-2, LMCI-1, and LMCI-2. Based on the relationship with the cognitive measures, trajectory analysis and the longitudinal dementia outcome of these patients, the progression of severity in the order of EMCI-1, EMCI-2, LMCI-1, and LMCI-2 is validated. Moreover, all four subtypes, as well as EMCI and LMCI classification, were all validated by the EFIGA dataset, through the metabolomics based label transfer, confirming findings obtained from the ADNI cohort.

At the molecular level, these subtypes differ significantly based on gene expression, metabolite intensity, and biological pathways.

The EMCI group and the LMCI group share several dysregulated metabolomic pathways, including alanine, aspartate, glutathione metabolism and glutamate metabolism, and arginine biosynthesis which are downregulated in both EMCI-2 and LMCI-2 subtypes. The consistent trend in the EMCI group and the LMCI group suggests these pathway changes may exist on a continuum as the disease progresses, similar to those reported in other studies ^44,45^. Among these pathways, disruptions in alanine, aspartate, and glutamate metabolism have been observed in ApoE4-carrying neuroglioma cells, indicating a connection between ApoE4 status and the metabolic dysregulation associated with AD progression ^46^. The downregulation of glutathione metabolism may contribute to the brain’s compromised antioxidant defense mechanism in the face of high oxidative stress. This aligns with the well-established previous finding that the concentration of glutathione decreases in the blood samples of AD patients compared to non-AD patients, suggesting a systemic reduction in antioxidant capacity ^47^. Additionally, disturbances in lipid metabolism, including glycerophospholipid metabolism have been reported in ApoE4 carriers, consistent with the observed upregulation of glycerophospholipid metabolism ^48^.

There also exist metabolic pathways that are not shared between EMCI and LMCI or dysregulated but towards different directions. Remarkably, tryptophan metabolism is upregulated in EMCI-2 but not enriched in LMCI-2. Tryptophan metabolism via the kynurenine pathway, driven by IDO1 activation, has been shown to impair astrocytic glycolysis and reduce neuronal energy support, contributing to synaptic dysfunction and cognitive decline ^49^. This suggests that the early upregulation of tryptophan metabolism in EMCI may mark the onset of metabolic dysfunction, while its absence in LMCI indicates that astrocytic support has already deteriorated by the later stages of the disease. The downregulation of arginine and proline metabolism, unique to the LMCI group, is in line with a previous finding that the arginine level in the temporal cortex is 27% lower in AD patients than in age-matched controls ^50^.

In the EMCI group, DNA damage response is active. Oxidative DNA damage has been found to occur in the early stage of AD progression ^51^. DNA repair is a consequential activity in the face of DNA damage. However, it is not clear yet what the exact cause of oxidative DNA damage is, and hypotheses include aging, exposure to neurotoxins, and early-life epigenetic alterations ^52^. In addition to DNA damage response, myeloid cell differentiation is suppressed in the EMCI group.

Myeloid-derived suppressor cells (MDSCs) are immature myeloid cells that exhibit robust suppressive function on T cell proliferation and mature myeloid cell function. Importantly, increased MDSCs in the peripheral immune system have been shown to enhance proinflammatory gene expression, contributing to a shift toward a proinflammatory immune environment even before substantial neurodegeneration occurs. This process involves the altered maturation and reduced functional capacity of monocytes and macrophages, potentially connecting the early immune response to the progression of neurodegenerative changes in AD ^53^.

The LMCI group has a marked shift towards calcium-related processes, with the significant activation of calcium channel regulator activity and calcium ion transport pathways. This calcium-related activation may reflect compensatory mechanisms or dysregulation of calcium homeostasis in later stages of cognitive impairment, such as increased calcium influx through NMDARs and RyRs, which can lead to synaptic dysfunction, mitochondrial stress, and tau hyperphosphorylation, ultimately impairing cognitive function and accelerating AD progression ^54^. The suppression of various metabolic processes, particularly those involved in DNA-templated transcription and organic acid metabolism, suggests a broader metabolic dysfunction as the condition progresses, such as impaired glycolysis and reduced monocarboxylic acid utilization, which limit neuronal energy supply and exacerbate oxidative stress, ultimately impairing synaptic function and accelerating cognitive decline ^55^.

The difference in activated pathways between the EMCI group and the LMCI group aligns with the well-established progression of Alzheimer’s disease, which begins with early oxidative stress and is further exacerbated by tau hyperphosphorylation ^56^. Our study pinpoints specific pathway changes at each stage of disease progression, distinguishing between early compensatory mechanisms in EMCI and metabolic dysfunction in LMCI. Notably, while some dysregulated pathways are shared across both stages, unique changes in each stage may help guide further investigation of AD biomarkers.

While significant efforts have been made to investigate MCI and its relationship to AD, most studies have relied on single-omics approaches or non-omics modalities. Multi-omics integration has emerged as a powerful strategy to refine disease classification by capturing the molecular complexity of MCI heterogeneity, yet relatively few studies have fully leveraged transcriptomics, metabolomics, or proteomics for this purpose ^19,57^. For example, previous research has demonstrated that integrating proteomics, lipidomics, and metabolomics significantly improved disease stratification, with such an approach achieving an Area Under the Curve (AUC) of 0.879, while lipidomic and metabolomic markers provided complementary insights into disease progression ^58^. Additionally, the combination of four molecules identified in the literature through multi-omics analysis improved the prediction of both AD status and cognitive decline, further supporting the effectiveness of integrating multiple biological layers ^59^.

Despite these advancements, current multi-omics studies have primarily focused on enhancing overall classification accuracy between AD, MCI, and normal patients, rather than identifying biologically distinct subtypes within MCI itself. Integrating multiple data modalities has proven to be effective in both prediction and classifications ^60^. Transcriptomics has been particularly useful in identifying disease-specific gene networks in AD, revealing key pathways associated with disease progression ^61^. MCI is a highly heterogeneous condition, and refining its subtypes is crucial for understanding disease progression, identifying high-risk individuals, and developing targeted early interventions ^62^. Our study uniquely addresses this gap by applying transcriptomics-metabolomics integration to refine MCI classification. Unlike previous studies, which have primarily focused on improving MCI classification at the population level ^63,64^, our approach identifies biologically distinct subgroups with unique metabolic and transcriptional signatures using SNF. These subtypes display differentially dysregulated metabolic and immune pathways, correlating with cognitive declining trajectories, longitudinal dementia risk, and biomarker validation (p-Tau181, p-Tau217, Aβ40, Aβ42 levels in EFIGA dataset). This novel stratification framework provides a biologically grounded refinement of MCI heterogeneity, a new perspective on MCI heterogeneity that could refine clinical stratification and advance precision preventative medicine.

There are some limitations worth mentioning in our current work. Most notably, even though the ADNI consortium represents a large-scale collaborative effort and contains a comprehensive database of elderly patients, the population is overwhelmingly white (92% in EMCI, 94.5% in LMCI). Confirmation of the observations to other ethnicities and racial groups should be followed up. Nevertheless, our study revealed subtypes in the EMCI and LMCI groups and identified the critical role of oxidative stress and metabolic pathways associated with progression in the pre-AD continuum. Such knowledge will help guide more effective drug development tailored towards subpopulations of patients in the MCI stage.

## 5. CONCLUSION

This study identified and validated distinct molecular subtypes within the early and late mild cognitive impairment (EMCI and LMCI) stages of Alzheimer’s disease, demonstrating that metabolomics plays a dominant role in subtype differentiation. The observed metabolic and transcriptomic differences align with disease progression and highlight distinct pathways involved in early oxidative stress responses and later metabolic dysfunction. These findings provide a refined molecular framework for MCI heterogeneity, supporting the development of precision medicine approaches and targeted interventions for individuals at risk of Alzheimer’s disease progression.

## Data Availability

All data produced in the present study are available upon reasonable request to the authors

## AUTHORS’ CONTRIBUTIONS

LG envisioned this project and supervised the study. BL did label transfer and subtype validation and wrote part of the manuscript. SY performed the initial data analysis on gene expression-metabolomics integration and wrote part of the initial manuscript. YD performed data analysis and revised the manuscript. SZ did the differential gene expression analysis and the time-to-event analysis, made the code available on the Github repository and revised the manuscript. LQ did the differential metabolomics analysis and improved the enrichment analysis. YZ pre-processed the ADNI metabolomics data. ZK prepared the data for the initial analysis and wrote part of the initial manuscript. LS provided feedback on the study design, result analysis and manuscript preparation. AL performed data analysis and contributed to manuscript writing in the EFIGA study. JL conducted data analysis in EFIGA. BV supervised the study in EFIGA.

## CONFLICTS OF INTEREST

The authors report no conflicts of interest.

## ACKNOWLEDGEMENT

We thank the Alzheimer’s Disease Neuroimaging Initiative (ADNI) for providing data for this research project. This study was supported by the National Institutes of Health grants R01 LM012373 and LM012907 from the National Library of Medicine (NLM), and grant R01 HD084633 from the Eunice Kennedy Shriver National Institute of Child Health and Human Development (NICHD), awarded to L.X. Garmire; as well as grants R01 AG071470 and U19 AG074879 from the National Institute on Aging (NIA) awarded to L. Shen.

Yuheng Du was partially supported by the Advanced Proteogenomics of Cancer Training Grant (T32 CA140044). Additionally, A.J.L. was supported by NIH grant K01 AG084849, NIH grant U19 AG078109, NIH grant UR013373, as well as funding from the Carol and Larry Ruvo Center for Brain Health through the National Alzheimer’s Coordinating Center and the Alzheimer’s Association.

## CODE AVAILABILITY

The code for this project is written in R and Python, and available at https://github.com/lanagarmire/AD_multi_Omics

---

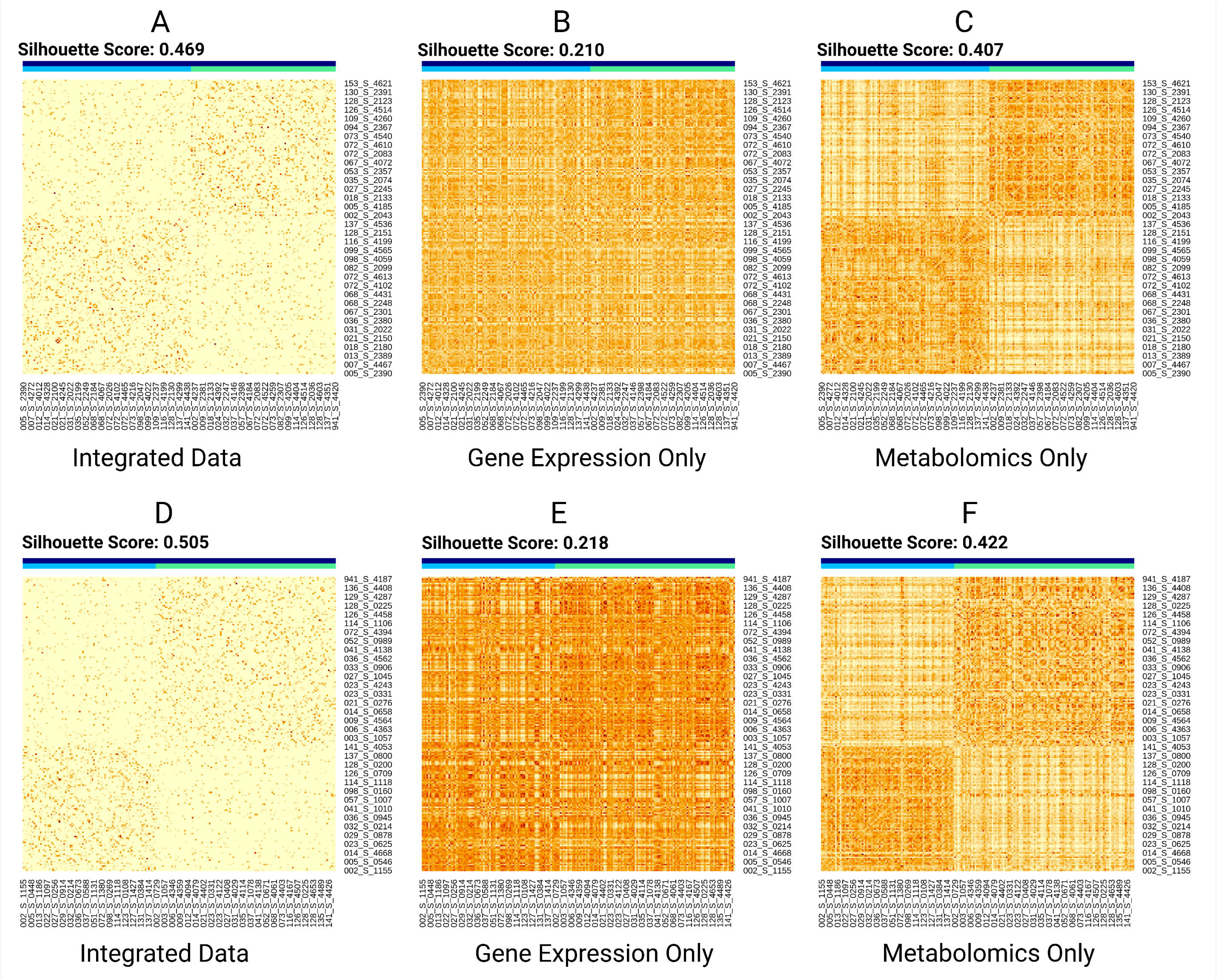

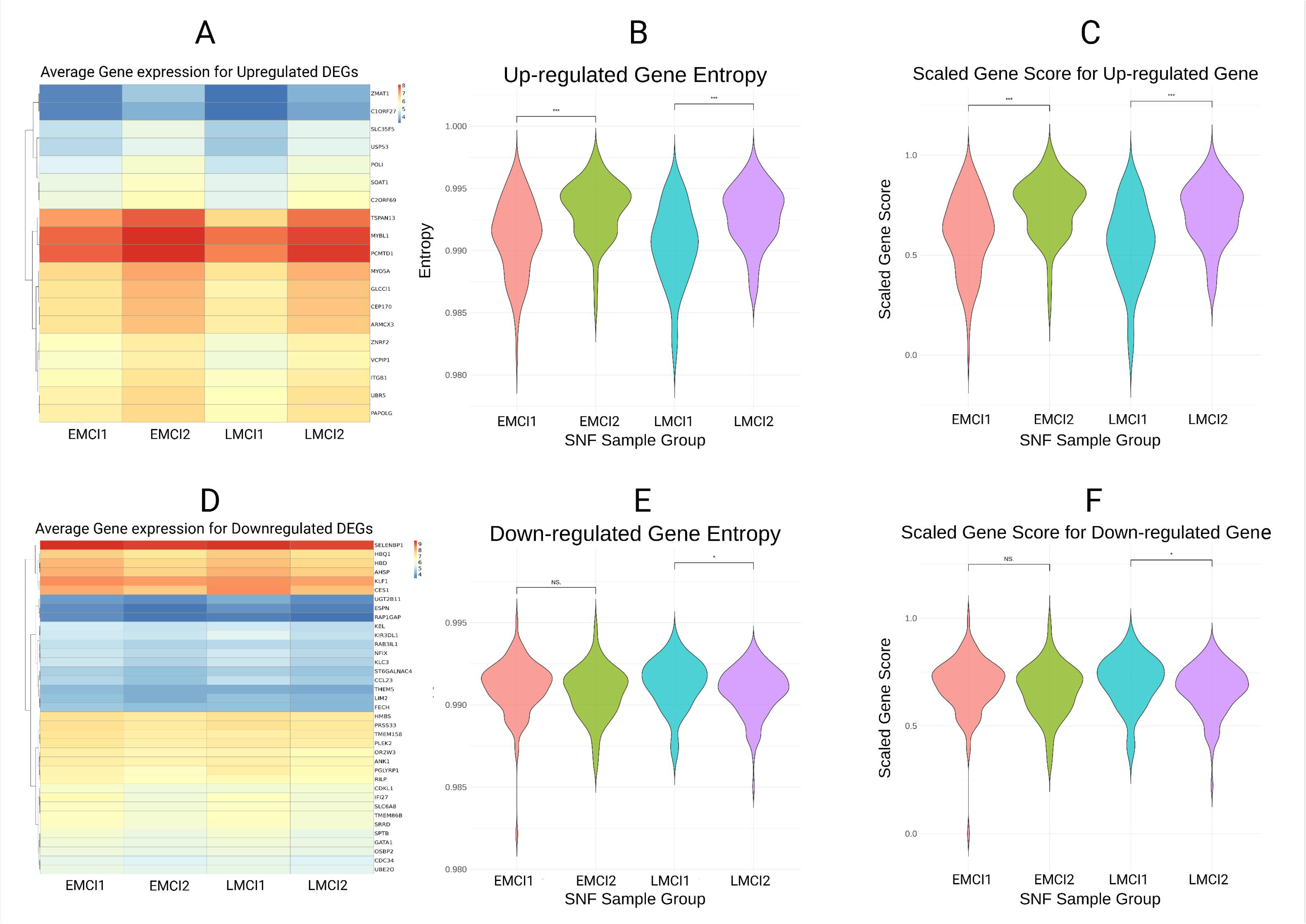

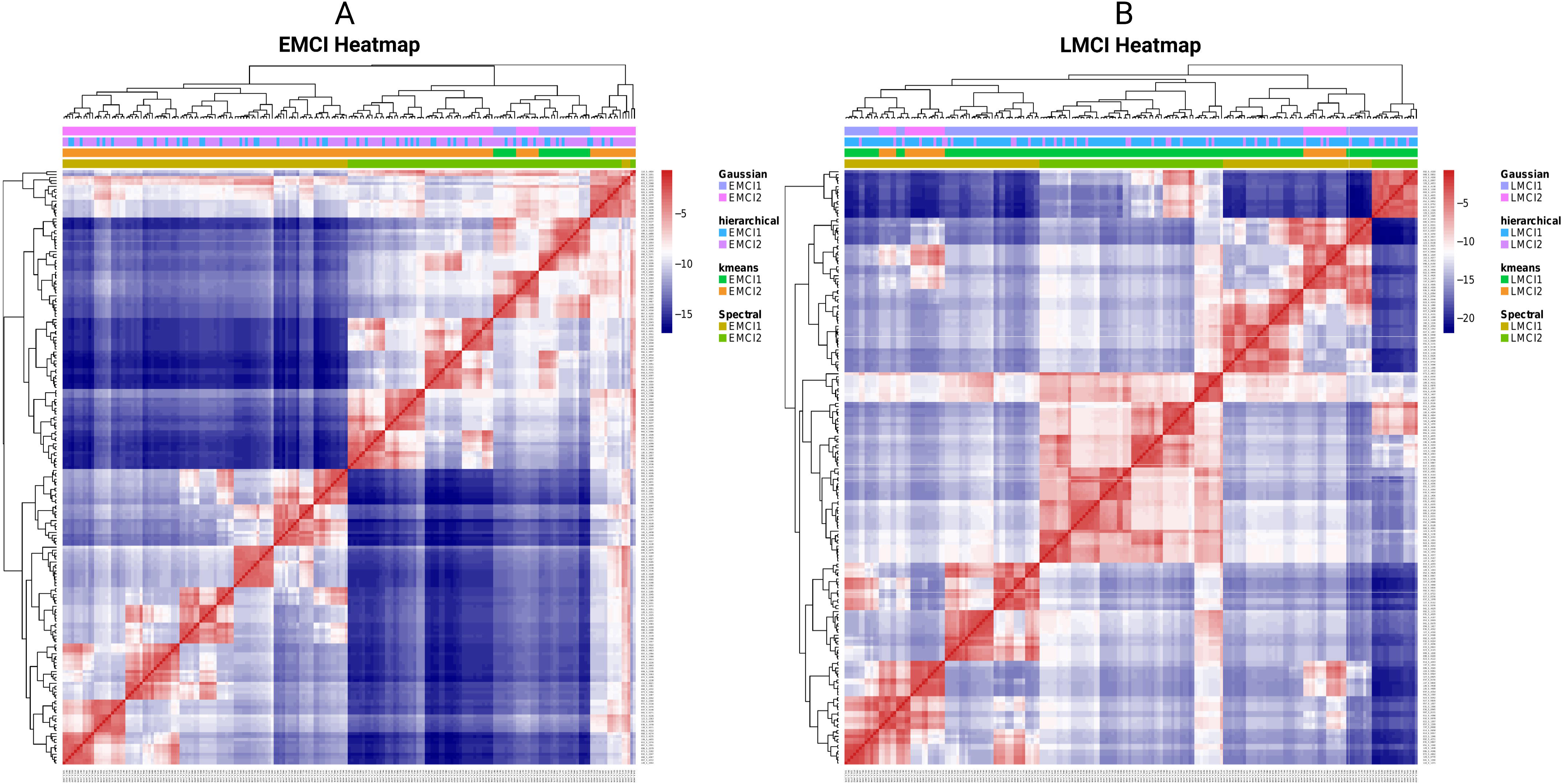

## Notes

### Competing Interest Statement

The authors have declared no competing interest.

### Funding Statement

This study was supported by the National Institutes of Health (NIH) grants R01 LM012373 and R01 LM012907 from the National Library of Medicine (NLM), and R01 HD084633 from the Eunice Kennedy Shriver National Institute of Child Health and Human Development (NICHD), awarded to L.X. Garmire. It was also supported by NIH grants R01 AG071470 and U19 AG074879 from the National Institute on Aging (NIA), awarded to L. Shen.
Yuheng Du was partially supported by the Advanced Proteogenomics of Cancer Training Grant (T32 CA140044).
A.J.L. was supported by NIH grant K01 AG084849, NIH grant U19 AG078109, NIH grant UR013373, as well as funding from the Carol and Larry Ruvo Center for Brain Health through the National Alzheimer's Coordinating Center and the Alzheimer's Association.

### Author Declarations

The ADNI Dataset

### Summary of Updates

We incorporated an external validation dataset from Columbia University and updated the results accordingly. Figures 4 to 6 were added to present the results from the validation. Figures 2 and 5 were revised. The Methods section was expanded with more details on preprocessing and integration. Funding information was also added.

## REFERENCES

1. Smoller, E. S. I Can’t Remember: Family Stories of Alzheimer’s Disease. (Temple University Press, 2010).

2. Frank, E. M. Effect of Alzheimer’s disease on communication function. J. S. C. Med. Assoc. 90, 417–423 (1994).

3. Ballard, C. et al. Alzheimer’s disease. Lancet 377, 1019–1031 (2011).

4. Prince, M. et al. The global prevalence of dementia: a systematic review and metaanalysis. Alzheimers. Dement. 9, 63–75.e2 (2013).

5. Jia, J. et al. The cost of Alzheimer’s disease in China and re-estimation of costs worldwide. Alzheimers. Dement. 14, 483–491 (2018).

6. Cisbani, G. & Rivest, S. Targeting innate immunity to protect and cure Alzheimer’s disease: opportunities and pitfalls. Mol. Psychiatry 26, 5504–5515 (2021).

7. Yiannopoulou, K. G. & Papageorgiou, S. G. Current and future treatments for Alzheimer’s disease. Ther. Adv. Neurol. Disord. 6, 19–33 (2013).

8. Petersen, R. C. Clinical subtypes of Alzheimer’s disease. Dement. Geriatr. Cogn. Disord. 9 Suppl 3, 16–24 (1998).

9. Edmonds, E. C. et al. Early versus late MCI: Improved MCI staging using a neuropsychological approach. Alzheimers. Dement. 15, 699–708 (2019).

10. Ströhle, A. et al. Drug and Exercise Treatment of Alzheimer Disease and Mild Cognitive Impairment: A Systematic Review and Meta-Analysis of Effects on Cognition in Randomized Controlled Trials. Am. J. Geriatr. Psychiatry 23, 1234–1249 (2015).

11. Cummings, J. L., Doody, R. & Clark, C. Disease-modifying therapies for Alzheimer disease: challenges to early intervention. Neurology 69, 1622–1634 (2007).

12. Olazarán, J. et al. Benefits of cognitive-motor intervention in MCI and mild to moderate Alzheimer disease. Neurology 63, 2348–2353 (2004).

13. Horr, T., Messinger-Rapport, B. & Pillai, J. A. Systematic review of strengths and limitations of randomized controlled trials for non-pharmacological interventions in mild cognitive impairment: focus on Alzheimer’s disease. J. Nutr. Health Aging 19, 141–153 (2015).

14. Mitelpunkt, A. et al. Novel Alzheimer’s disease subtypes identified using a data and knowledge driven strategy. Sci. Rep. 10, 1327 (2020).

15. Dettmer, K. & Aronov, P. A. Mass spectrometry-based metabolomics. spectrometry reviews (2007).

16. Molecular Characterization and Clinical Relevance of Metabolic Expression Subtypes in Human Cancers. Cell Reports 23, 255–269.e4 (2018).

17. Gao, G. F. et al. Before and After: Comparison of Legacy and Harmonized TCGA Genomic Data Commons’ Data. Cell Syst 9, 24–34.e10 (2019).

18. Wilkins, J. M. & Trushina, E. Application of Metabolomics in Alzheimer’s Disease. Front. Neurol. 8, 719 (2017).

19. Badhwar, A. et al. A multiomics approach to heterogeneity in Alzheimer’s disease: focused review and roadmap. Brain 143, 1315–1331 (2020).

20. Yan, Z. et al. Presymptomatic Increase of an Extracellular RNA in Blood Plasma Associates with the Development of Alzheimer’s Disease. Curr Biol 30, 1771–1782.e3 (2020).

21. Liu, F., Wee, C.-Y., Chen, H. & Shen, D. Inter-modality relationship constrained multi-modality multi-task feature selection for Alzheimer’s Disease and mild cognitive impairment identification. Neuroimage 84, 466–475 (2014).

22. Wang, B. et al. Similarity network fusion for aggregating data types on a genomic scale. Nat. Methods 11, 333–337 (2014).

23. Petersen, R. C. et al. Alzheimer’s Disease Neuroimaging Initiative (ADNI): clinical characterization. Neurology 74, 201–209 (2010).

24. Mueller, S. G. et al. Ways toward an early diagnosis in Alzheimer’s disease: the Alzheimer’s Disease Neuroimaging Initiative (ADNI). Alzheimers Dement 1, 55–66 (2005).

25. St John-Williams, L., et al. Bile acids targeted metabolomics and medication classification data in the ADNI1 and ADNIGO/2 cohorts. Sci Data 6, 212 (2019).

26. Yasenko, L., Klyatchenko, Y. & Tarasenko-Klyatchenko, O. Image noise reduction by denoising autoencoder. in 2020 IEEE 11th International Conference on Dependable Systems, Services and Technologies (DESSERT) 351–355 (IEEE, 2020).

27. Nadel, B. B. et al. The Gene Expression Deconvolution Interactive Tool (GEDIT): accurate cell type quantification from gene expression data. Gigascience 10, (2021).

28. Newman, A. M. et al. Robust enumeration of cell subsets from tissue expression profiles. Nat. Methods 12, 453–457 (2015).

29. Pang, Z. et al. MetaboAnalyst 6.0: towards a unified platform for metabolomics data processing, analysis and interpretation. Nucleic Acids Res 52, W398–W406 (2024).

30. Gu, Z., Eils, R. & Schlesner, M. Complex heatmaps reveal patterns and correlations in multidimensional genomic data. Bioinformatics 32, 2847–2849 (2016).

31. Golovenkin, S. E. et al. Trajectories, bifurcations, and pseudo-time in large clinical datasets: applications to myocardial infarction and diabetes data. Gigascience 9, (2020).

32. He, B. et al. ASGARD is A Single-cell Guided Pipeline to Aid Repurposing of Drugs. Nat. Commun. 14, 993 (2023).

33. Alzheimer’s Investigator Assets. Columbia Neurology https://www.neurology.columbia.edu/research/research-centers-and-programs/alzheimers-disease-research-center-adrc/investigators (2020).

34. Jack, C. R. et al. NIA-AA Research Framework: Toward a biological definition of Alzheimer’s disease. Alzheimer’s & dementia : the journal of the Alzheimer’s Association 14, (2018).

35. Yu, T., Park, Y., Johnson, J. M. & Jones, D. P. apLCMS--adaptive processing of high-resolution LC/MS data. Bioinformatics 25, 1930–1936 (2009).

36. Uppal, K. et al. xMSanalyzer: automated pipeline for improved feature detection and downstream analysis of large-scale, non-targeted metabolomics data. BMC Bioinformatics 14, 15 (2013).

37. Honig, L. S. et al. Evaluation of Plasma Biomarkers for A/T/N Classification of Alzheimer Disease Among Adults of Caribbean Hispanic Ethnicity. JAMA Netw Open 6, e238214 (2023).

38. Cao, K., Bai, X., Hong, Y. & Wan, L. Unsupervised topological alignment for single-cell multi-omics integration. Bioinformatics 36, i48–i56 (2020).

39. Iqbal, M., Lee, C. K. M. & Keung, K. L. An intelligent fault diagnosis method based on L2 regularization and deep transfer LSTM. in 2024 IEEE International Conference on Industrial Engineering and Engineering Management (IEEM) 0282–0286 (IEEE, 2024).

40. Albergante, L. et al. Robust and Scalable Learning of Complex Intrinsic Dataset Geometry via ElPiGraph. Entropy 22, (2020).

41. Tissot, C. et al. Plasma pTau181 predicts cortical brain atrophy in aging and Alzheimer’s disease. Alzheimers Res Ther 13, 69 (2021).

42. Core blood biomarkers of Alzheimer’s disease: A single-center real-world performance study. The Journal of Prevention of Alzheimer’s Disease 12, 100027 (2025).

43. Tong, J. et al. Poly-Adenine Assisted Signaling Displaced Probe Ratiometric Electrochemical Aptasensor for Accurate Detection of Alzheimer’s Disease Aβ Biomarkers. ACS Appl Mater Interfaces 16, 64297–64306 (2024).

44. Pietronigro, E., Zenaro, E. & Constantin, G. Imaging of Leukocyte Trafficking in Alzheimer’s Disease. Front. Immunol. 7, 33 (2016).

45. Yuan, Y., Niu, F., Liu, Y. & Lu, N. Zinc and its effects on oxidative stress in Alzheimer’s disease. Neurol. Sci. 35, 923–928 (2014).

46. Lu, L. et al. Metabolomic and Proteomic Analysis of ApoE4-Carrying H4 Neuroglioma Cells in Alzheimer’s Disease Using OrbiSIMS and LC-MS/MS. Anal Chem 96, 11760–11770 (2024).

47. Bermejo, P. et al. Peripheral levels of glutathione and protein oxidation as markers in the development of Alzheimer’s disease from Mild Cognitive Impairment. Free Radic Res 42, 162–170 (2008).

48. Chouinard-Watkins, R. & Plourde, M. Fatty acid metabolism in carriers of apolipoprotein E epsilon 4 allele: is it contributing to higher risk of cognitive decline and coronary heart disease? Nutrients 6, 4452–4471 (2014).

49. Minhas, P. S. et al. Restoring hippocampal glucose metabolism rescues cognition across Alzheimer’s disease pathologies. Science 385, eabm6131 (2024).

50. Altiné-Samey, R. et al. The contributions of metabolomics in the discovery of new therapeutic targets in Alzheimer’s disease. Fundam. Clin. Pharmacol. 35, 582–594 (2021).

51. Lovell, M. A. & Markesbery, W. R. Oxidative DNA damage in mild cognitive impairment and late-stage Alzheimer’s disease. Nucleic Acids Res. 35, 7497–7504 (2007).

52. Coppedè, F. & Migliore, L. DNA damage in neurodegenerative diseases. Mutat. Res. 776, 84–97 (2015).

53. Thome, A. D. et al. Functional alterations of myeloid cells during the course of Alzheimer’s disease. Molecular Neurodegeneration 13, 1–11 (2018).

54. Ge, M. et al. Role of Calcium Homeostasis in Alzheimer’s Disease. Neuropsychiatr Dis Treat 18, 487–498 (2022).

55. APOE and metabolic dysfunction in Alzheimer’s disease. in International Review of Neurobiology vol. 154 131–151 (Academic Press, 2020).

56. Iatrou, A., Clark, E. M. & Wang, Y. Nuclear dynamics and stress responses in Alzheimer’s disease. Mol. Neurodegener. 16, 65 (2021).

57. Huang, S., Chaudhary, K. & Garmire, L. X. More Is Better: Recent Progress in Multi-Omics Data Integration Methods. Front. Genet. 8, 268903 (2017).

58. Filippos Anagnostakis,Michail Kokkorakis, Keenan A. Walker and Christos Davatzikos. Signatures and Discriminative Abilities of Multi-Omics between States of Cognitive Decline. Biomedicine (2024) doi:10.3390/biomedicines12050941.

59. Clark, C., Dayon, L., Masoodi, M., Bowman, G. L. & Popp, J. An integrative multi-omics approach reveals new central nervous system pathway alterations in Alzheimer’s disease. Alzheimers Res Ther 13, 71 (2021).

60. Zhang, S. et al. PheME: A deep ensemble framework for improving phenotype prediction from multi-modal data. in 2023 IEEE 11th International Conference on Healthcare Informatics (ICHI) 268–275 (IEEE, 2023).

61. Xiang, S. et al. Condition-specific gene co-expression network mining identifies key pathways and regulators in the brain tissue of Alzheimer’s disease patients. BMC Medical Genomics 11, 39–51 (2018).

62. Anderson, N. D. State of the science on mild cognitive impairment (MCI). CNS Spectr 24, 78–87 (2019).

63. Feng, Y. et al. Deep multiview learning to identify imaging-driven subtypes in mild cognitive impairment. BMC Bioinformatics 23, 402 (2022).

64. Yuan, S., Li, H., Wu, J. & Sun, X. Classification of Mild Cognitive Impairment With Multimodal Data Using Both Labeled and Unlabeled Samples. IEEE/ACM Trans Comput Biol Bioinform 18, 2281–2290 (2021).

