## Supplementary Document for "Identification and Validation of New Molecular Subtypes within the Early and Late Mild Cognitive Impairment Stages of Alzheimer’s Disease"

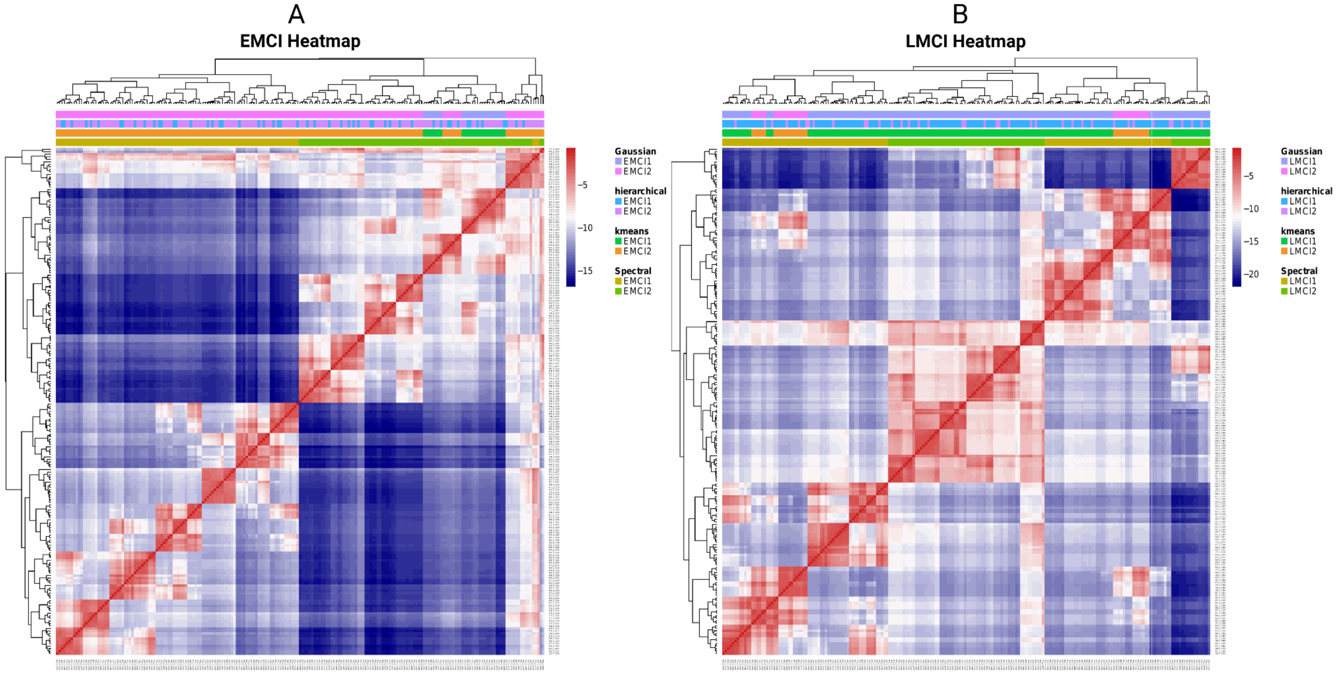


**Supplementary Figure 1.** Heatmaps showing clustering results for early mild cognitive impairment (EMCI) and late mild cognitive impairment (LMCI) subgroups using four clustering methods: Gaussian mixture models, hierarchical clustering, k-means, and spectral clustering. Spectral clustering demonstrated the highest consistency and distinction among the four methods. The subgroups (EMCI1, EMCI2, LMCI1, and LMCI2) were distinguished based on the clustering patterns and annotations derived from the predicted labels of each method. (A) EMCI1 and EMCI2 subgroups. (B) LMCI1 and LMCI2 subgroups.

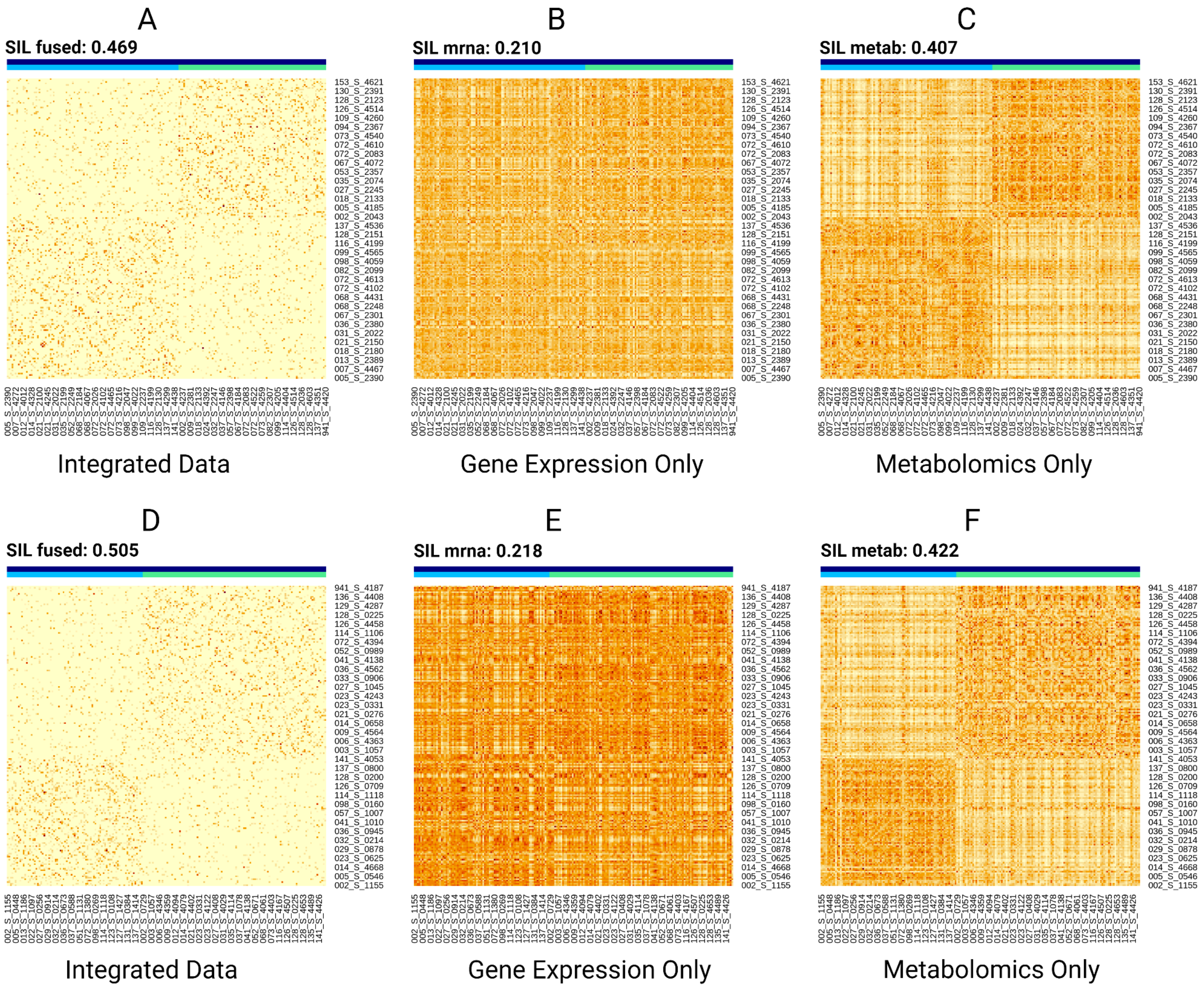


**Supplementary Figure 2.** Visualization of patient-level spectral clustering results for the two clusters, EMCI and LMCI, using integrated data, gene expression only, and metabolomics only. Panels A–C show clustering results for EMCI, while panels D–F show results for LMCI. The Silhouette Scores (SIL) indicate that metabolomics data play a more significant role in distinguishing subtypes compared to gene expression data.

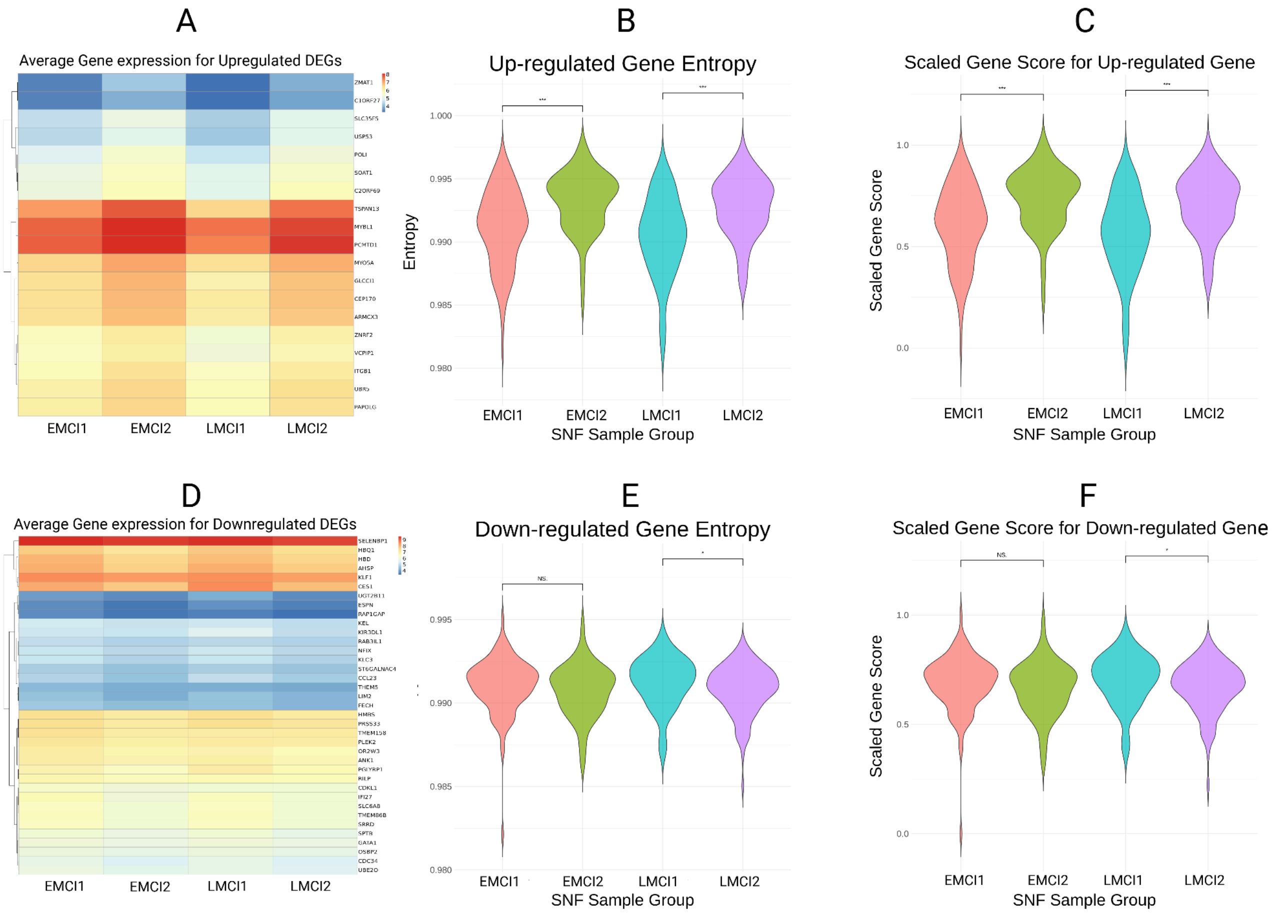


**Supplementary Figure 3.** Comparison of average gene expression for up-regulated and down-regulated differentially expressed genes (DEGs) among EMCI and LMCI subtypes. Analysis of gene expression patterns revealed distinct differences between subtypes. For up-regulated DEGs, EMCI-2 exhibited higher average expression compared to EMCI-1, and LMCI-2 showed higher expression compared to LMCI-1, as visualized in (A). For down-regulated DEGs (D), LMCI-2 demonstrated significantly lower expression than LMCI-1, while differences between EMCI-1 and EMCI-2 were statistically insignificant. The statistical significance of these expression differences was further validated through analysis of gene entropy (violin plots in B and E) and scaled gene scores (C and F). Both measures indicated significantly higher scores in the later stages for EMCI and LMCI subtypes, except for the down-regulated genes in EMCI-1 and EMCI-2. These findings confirm the molecular distinctions between the subtypes based on gene expression levels and derived scores.
